# The Impact of Digital Health on Maternal Mental Health in Nigeria

**DOI:** 10.1101/2025.04.02.25324872

**Authors:** Oluwafemi Babatunde Banjo

**Author notes:** Corresponding author: Oluwafemi B. Banjo.

## Abstract

Maternal mental health remains a critical yet underrecognized public health concern in Nigeria, with postpartum depression (PPD) and maternal suicide emerging as significant challenges due to limited access to mental healthcare services, stigma, and systemic barriers. This study explores the potential of digital health solutions such as mobile applications, teletherapy, AI-driven mental health chatbots, and SMS-based interventions in addressing postpartum mental health disorders among Nigerian mothers. Using a quantitative survey of 200 postpartum women, the research examines the effectiveness, accessibility, and challenges associated with digital mental health interventions.

## 1. Introduction

### 1.1 Background of Maternal Mental Health in Nigeria

Maternal mental health is an essential yet often overlooked component of maternal healthcare in Nigeria. Pregnancy and childbirth bring significant physiological, psychological, and social changes, which, if not adequately managed, can lead to adverse mental health outcomes.

Among the most common maternal mental health disorders is postpartum depression (PPD), which affects an estimated 10–20% of Nigerian mothers, with a higher prevalence in rural and underserved communities (Adewuya et al., 2022). Other conditions, such as postpartum anxiety, post-traumatic stress disorder (PTSD), and maternal suicide ideation, also contribute to the growing public health burden associated with maternal mental health disorders.

Despite the prevalence of maternal mental health challenges, the issue remains largely underreported and inadequately addressed. The stigma surrounding mental illness in Nigeria discourages affected women from seeking professional help, leading many to suffer in silence. Cultural perceptions often associate postpartum distress with spiritual or supernatural causes rather than treatable medical conditions, further exacerbating barriers to timely intervention.

Additionally, gender roles and societal expectations place a high burden on Nigerian mothers, with many experiencing intense psychological stress due to financial insecurity, domestic violence, and lack of spousal or family support.

Nigeria’s healthcare system also presents significant structural barriers to maternal mental healthcare. Mental health services are limited, with fewer than 300 psychiatrists serving a population exceeding 200 million (World Health Organization, 2023). In many primary healthcare facilities, maternal mental health screenings are not integrated into routine postnatal care, leaving many cases undiagnosed. Moreover, women in rural and underserved areas have limited access to specialized care due to a shortage of trained mental health professionals and inadequate infrastructure.

The burden of maternal suicide is another critical concern. Although official statistics on maternal suicide rates in Nigeria remain scarce due to stigma and misclassification of deaths, existing studies indicate that maternal suicide is a rising yet underrecognized problem (Ogunsemi et al., 2021). The interplay of postpartum depression, economic hardship, intimate partner violence, and societal pressures increases the risk of suicide among vulnerable mothers.

In recent years, digital health solutions have emerged as a potential avenue for improving maternal mental healthcare access and outcomes. Mobile health (mHealth) applications, telepsychiatry, and online peer support platforms offer innovative approaches to bridging the gap between affected mothers and mental health professionals. However, despite their promise, these solutions face challenges such as digital illiteracy, high internet costs, data privacy concerns, and low adoption rates due to prevailing cultural beliefs.

Addressing maternal mental health in Nigeria requires a multi-sectoral approach that integrates digital health innovations, public health policies, community-based interventions, and increased awareness campaigns to reduce stigma. Strengthening healthcare infrastructure, expanding mental health training for healthcare workers, and promoting the use of digital mental health services can significantly enhance support for postpartum women and mitigate the long-term impact of maternal mental disorders on families and society.

### 1.2 Digital Health and Its Role in Mental Healthcare

The integration of digital health technologies into mental healthcare has revolutionized the accessibility and delivery of psychological support services worldwide. In Nigeria, where maternal mental health services are often scarce or inaccessible, digital health solutions provide an innovative and scalable approach to addressing postpartum depression (PPD) and related conditions. These solutions leverage mobile technology, telemedicine, artificial intelligence (AI), and digital platforms to bridge the gap between healthcare providers and mothers in need of support.

One of the primary benefits of digital health interventions is their ability to overcome geographical and systemic barriers to mental healthcare. Mobile health (mHealth) applications offer self-assessment tools, guided cognitive behavioural therapy (CBT), and symptom-tracking features that enable mothers to monitor their mental well-being. Examples include mobile-based mental health screening tools that help detect early symptoms of PPD and provide immediate recommendations for intervention (Naslund et al., 2020).

Teletherapy and virtual counselling platforms have also gained traction, allowing postpartum women to access professional psychological support remotely. Through video calls, voice consultations, or text-based therapy sessions, mothers can engage with mental health professionals without the logistical and financial constraints of in-person visits. This is particularly crucial in rural and underserved communities, where psychiatric services are often unavailable or underfunded.

AI-powered mental health chatbots present another innovative digital intervention. These chatbots, designed with natural language processing capabilities, provide real-time emotional support, psychoeducation, and coping strategies for postpartum women experiencing distress. AI-driven platforms can assess mood patterns, suggest mental health resources, and escalate cases to human therapists when necessary, ensuring timely intervention (Torous et al., 2021).

Furthermore, SMS-based mental health support systems have been successfully implemented in low-resource settings to provide mothers with mental health education, stress management tips, and reminders for postnatal care appointments. Given the widespread use of mobile phones across Nigeria, even in low-income communities, SMS-based interventions can serve as an effective tool for reaching a broader population of postpartum women.

### 1.3 Rationale for the Study

Despite the increasing adoption of digital health technologies in Nigeria, research on their effectiveness in addressing maternal mental health remains limited. Postpartum depression (PPD) and maternal suicide are significant public health concerns, yet access to mental health services remains inadequate due to stigma, financial constraints, and a shortage of trained professionals. Digital health interventions, including mobile applications, teletherapy, and AI-powered mental health tools, offer a promising avenue for bridging this gap.

This study seeks to evaluate the extent to which digital health solutions can improve maternal mental health outcomes by providing timely, accessible, and cost-effective interventions that will ultimately enhance maternal well-being in Nigeria.

### 1.4 Research Objectives

1. Assess the effectiveness of mobile mental health solutions in managing postpartum depression.
2. Identify the barriers to digital mental health adoption among Nigerian mothers.
3. Explore the role of digital interventions in rural communities.
4. Provide policy recommendations to improve digital maternal mental healthcare.

### 1.5 Scope and Limitations

This study focuses on postpartum mothers across Nigeria, utilizing data from 200 survey respondents to assess the impact of digital health solutions on maternal mental health. While the findings provide valuable insights with regard to the information at the disposal of these mothers, the results may not be fully representative of all Nigerian states due to the number of mothers surveyed. Nonetheless, the study offers a crucial foundation for future research and policy development in digital maternal mental healthcare.

## 2. Literature Review

### 2.1 Maternal Mental Health: Global and Nigerian Perspectives

Maternal mental health is a significant public health concern worldwide, with postpartum depression (PPD) affecting approximately 10–15% of women globally (WHO, 2023). The burden of PPD is even more pronounced in low- and middle-income countries, where socio-economic disparities, inadequate healthcare infrastructure, and cultural stigma contribute to higher prevalence rates. In Nigeria, studies estimate that between 14% and 30% of postpartum women experience depressive symptoms, with higher rates observed in rural and underserved communities due to limited access to mental health services, financial stress, and weak social support systems.

Despite the growing recognition of maternal mental health as a critical component of overall well-being, traditional postpartum care in Nigeria predominantly focuses on physical recovery and infant health, often neglecting the psychological well-being of mothers. Cultural norms and societal expectations frequently discourage open discussions about mental health, leading to delayed or missed interventions. As a result, many women suffer in silence, increasing the risk of severe mental health conditions, including maternal suicide.

### 2.2 Prevalence and Risk Factors of Postpartum Depression in Nigeria

Postpartum depression (PPD) is a significant public health concern in Nigeria, with prevalence rates varying across regions due to socio-economic and cultural factors. Data from 200 postpartum mothers in Nigeria indicate that 22% reported symptoms of moderate to severe depression, aligning with national estimates. However, this figure may be an underrepresentation due to the stigma surrounding mental health, which often discourages affected women from seeking help or acknowledging their symptoms.

Several risk factors contribute to the high prevalence of PPD in Nigeria. Financial stress remains one of the leading triggers, particularly among low-income families where the burden of childcare expenses exacerbates emotional distress. Additionally, the lack of spousal support significantly influences maternal mental health, as many Nigerian women face the pressure of managing postpartum recovery, childcare responsibilities, and household duties with minimal assistance. Cultural stigma further compounds these challenges, as mental health issues are often dismissed or misunderstood within traditional settings, leading to social isolation and delayed intervention.

Beyond these primary factors, other contributors to PPD include hormonal changes, past trauma, pregnancy complications, and the psychological impact of unplanned pregnancies. In rural and underserved communities, limited access to maternal healthcare services and professional mental health support further escalates the risk.

### 2.3 Maternal Suicide: An Emerging Public Health Concern

Maternal suicide, though often underreported in Nigeria, is an increasing public health concern, particularly among women experiencing severe, untreated postpartum depression. The combination of social stigma, lack of mental health resources, and inadequate screening in maternal care settings leave many cases undetected until crisis points are reached. Limited awareness of digital mental health solutions further hinders early intervention, reducing opportunities for timely psychological support. Addressing this issue requires greater integration of mental health screening into maternal healthcare services and the promotion of accessible digital interventions to provide immediate and confidential support.

### 2.4 Digital Health Solutions in Mental Healthcare: Global Best Practices

The integration of digital health solutions into mental healthcare has shown promising results globally, particularly in low- and middle-income countries (LMICs). In Kenya and India, targeted mobile mental health interventions have demonstrated up to a 40% reduction in depression rates among users (Ochieng et al., 2022). These interventions include smartphone applications for mood tracking, AI-powered chatbots for psychological support, and teletherapy platforms connecting patients with mental health professionals.

For Nigeria, replicating these models within the maternal healthcare system presents a viable approach to addressing postpartum depression and maternal suicide. Mobile applications tailored for postpartum mothers could provide real-time emotional support. Additionally, SMS-based counselling services for low-literacy and rural populations could enhance accessibility.

To maximize impact, Nigeria must prioritize government-led digital health policies, public-private collaborations, and community engagement strategies that promote digital mental health literacy among mothers.

### 2.5 Mobile Mental Health Interventions: Effectiveness and Challenges

Mobile mental health interventions have shown promise in addressing postpartum depression by providing accessible and cost-effective psychological support. In several countries, AI-powered mental health chatbots and teletherapy platforms have demonstrated success in reducing depression rates among postpartum women. These digital tools offer immediate, anonymous, and stigma-free support, allowing mothers to access counselling, guided self-help resources, and crisis intervention services without the need for in-person visits.

Despite their potential, several challenges hinder the widespread adoption of mobile mental health solutions in Nigeria. Digital literacy remains a major barrier, particularly in rural communities where access to smartphones and internet connectivity is limited. Additionally, the cultural stigma surrounding mental health continues to discourage many women from seeking help, even when digital interventions are available. Affordability is another critical issue, as many postpartum women in low-income households struggle to afford mobile data, smart devices, or subscription-based mental health services.

## 3. Methodology

### 3.1 Research Design

This study adopted a quantitative survey design to collect and analyze data on maternal mental health and digital health adoption among postpartum women in Nigeria. The survey method was chosen for its ability to capture measurable trends, identify patterns and understand people’s knowledge of digital health.

### 3.2 Study Population and Sampling Techniques

Data was collected from 200 postpartum mothers in Nigeria using randomized cluster sampling. The questionnaire and data retrieved are attached to the research.

### 3.3. Ethical Considerations in Digital Mental Health Research

Participants provided informed consent, and data confidentiality was maintained per the Nigeria Data Protection Regulation (NDPR). No personal details of the research will be disclosed.

## 4. Digital Health Solutions for Maternal Mental Health

### 4.1 Mobile Apps for Postpartum Depression Screening and Support

Digital health platforms have become essential tools for addressing postpartum depression (PPD) by providing accessible mental health support to new mothers. In Nigeria, mobile applications such as FriendnPal, PsyndUp, Truthshare, MyCareBuddy, Helpmum and others offer vital services, including PPD screening, self-help resources, and direct access to professional counselling. FriendnPal, recognized as Africa’s first AI-driven mental health platform, actively engages young people; particularly women through automated mental health assessments and personalized coping strategies.

Additionally, organizations like Mentally Aware Nigeria Initiative (MANI) provide digital follow-up checks on mothers experiencing mental health challenges, ensuring continuity of care. These mobile interventions not only bridge the gap in mental health service delivery but also help to reduce stigma by offering discreet and affordable support.

### 4.2 AI-Driven Chatbots and Mental Health Assistance

AI-driven chatbots are transforming maternal mental health care by providing immediate, anonymous, and 24/7 psychological support to postpartum women. Platforms like Pal, developed by FriendnPal, leverage artificial intelligence to offer real-time emotional assistance to women. These chatbots can communicate in local African languages, making them more accessible to diverse populations, particularly in underserved regions where traditional mental health services are limited.

Beyond chatbot support, these platforms also serve as a bridge to human-led interventions by connecting users with licensed therapists, mental health professionals, and peer support groups for further assistance.

To maximize the impact of AI-driven mental health solutions, hospitals and healthcare institutions should integrate digital screening tools into routine prenatal and postnatal care. By incorporating chatbot-based mental health assessments during maternal checkups, healthcare providers can identify at-risk women early, offer timely interventions, and reduce the long-term impact of postpartum depression and anxiety on both mothers and their infants.

## 5. Barriers to Adoption of Digital Mental Health Solutions

Despite the growing potential of digital mental health interventions, several structural and societal challenges hinder their widespread adoption among postpartum women in Nigeria. These barriers include digital literacy gaps, social stigma, infrastructural limitations, and data privacy concerns, all of which affect the accessibility and effectiveness of these solutions.

Digital Literacy: A significant proportion of postpartum women, particularly in rural and low-income communities, have limited experience with smartphones and digital applications. Many struggle with engaging mental health platforms, making it difficult for them to utilize digital interventions effectively. Without targeted digital literacy programs, these solutions may remain inaccessible to the most vulnerable populations.

Stigma: Mental health remains highly stigmatized in Nigeria, leading many women to avoid seeking help due to fear of judgment or discrimination. Cultural beliefs often equate mental health struggles with spiritual weakness or personal failure, making it difficult for postpartum women to acknowledge their symptoms or openly discuss their mental well-being. This stigma extends to digital platforms, where some women fear being identified as having mental health concerns.

Internet & Power Issues: Limited broadband access, unstable electricity supply, and high data costs pose major challenges, particularly in remote and underserved areas. Many women lack reliable internet connections or smartphone access, making it difficult for them to engage with online counselling, teletherapy, or AI-driven mental health services. The high cost of mobile data further restricts the continuous usage of digital mental health platforms.

Data Privacy Concerns: Privacy remains a key concern for many postpartum women when considering digital mental health solutions. Fear of data breaches, unauthorized access, or misuse of personal health information discourages women from fully utilizing these platforms. Without clear regulations and strict confidentiality assurances, concerns around digital mental health solutions may persist.

Addressing these barriers requires policy-driven interventions, increased awareness, and investment in digital infrastructure to ensure that these women can safely and effectively access mental health support through digital platforms.

## 6. The Role of Digital Health in Rural and Underserved Communities

### 6.1 Community-Based Mobile Mental Health Interventions

Community-driven digital health interventions are emerging as effective solutions for addressing postpartum depression (PPD) in Nigeria. In particular, mobile-based programs, such as SMS mental health interventions, have demonstrated measurable improvements in maternal mental health outcomes, especially in underserved regions.

A notable case study from Northern Nigeria by (Yusuf, A., & Adepoju, A. 2023) highlights the effectiveness of an SMS-based mental health support program designed for postpartum women in rural communities. This initiative provided structured psychoeducational messages, coping strategies, and reminders to encourage help-seeking behaviour. Participants reported a 15% reduction in PPD symptoms within three months of engagement. The program was particularly effective in reaching women who lacked access to conventional mental health services, demonstrating the potential of low-cost, scalable interventions in addressing maternal mental health challenges.

Despite these promising outcomes, barriers such as mobile phone ownership, literacy levels, and cultural stigma remain significant hurdles to adoption. Future interventions must integrate partnerships with community health workers to ensure accessibility and sustained engagement. Scaling such interventions nationwide could significantly improve mental health outcomes for postpartum women, particularly in low-resource settings where traditional healthcare access is limited.

## 7. Policy Recommendations and Future Directions

The integration of digital health solutions into maternal mental healthcare in Nigeria has shown promising potential. Both digital health platforms and government initiatives have made commendable efforts to expand access to mental health support. However, significant gaps remain, requiring targeted policy interventions and strategic investments to enhance effectiveness and sustainability.

Expand Teletherapy Services and Improve Digital Literacy:

Digital health platforms such as FriendnPal, PsyndUp, MyCareBuddy, Helpmum and Truthshare have been instrumental in providing remote therapy and mental health support for postpartum women. To scale their impact, government agencies and healthcare institutions should increase funding for teletherapy programs, subsidize digital mental health services, and integrate these platforms into public healthcare systems. Additionally, digital literacy training should be prioritized for postpartum women, ensuring they can effectively navigate mental health applications and online support systems.

Strengthen Data Protection Laws and AI-Based Mental Health Chatbots:

The adoption of AI-driven chatbots for maternal mental health, such as Pal by FriendnPal, has provided round-the-clock support to women experiencing postpartum depression. While these innovations are commendable, robust data protection policies must be enforced to safeguard user privacy and build trust in digital mental health services. The government should collaborate with digital health innovators to develop secure, ethical, and AI-driven mental health platforms that comply with global standards for data security and patient confidentiality.

To achieve widespread adoption of digital mental health solutions, multi-sectoral partnerships between policymakers, healthcare providers, tech innovators, and community organizations are crucial. By fostering an enabling environment for digital mental health integration, Nigeria can make significant strides in reducing postpartum depression rates, enhancing maternal well-being, and strengthening its overall mental healthcare system.

## 8. Conclusion

The integration of digital health solutions into maternal mental healthcare presents a promising avenue for addressing postpartum depression and reducing maternal suicide rates in Nigeria. The findings of this study confirm the critical gaps in traditional maternal mental health services, including limited accessibility, stigma, and inadequate healthcare infrastructure, which hinder timely intervention for postpartum women. The adoption of mobile mental health applications, teletherapy, AI-driven chatbots, and SMS-based support systems has demonstrated significant potential in bridging these gaps by providing accessible, cost-effective, and scalable interventions.

Despite the evident benefits, several barriers to the widespread adoption of digital mental health solutions persist. Issues such as digital illiteracy, social stigma, internet connectivity challenges, and data privacy concerns continue to limit the reach and effectiveness of these interventions, particularly in rural and underserved communities. Addressing these challenges requires a multi-faceted approach that includes policy-driven initiatives, increased investment in digital infrastructure, and targeted awareness campaigns to normalize discussions around maternal mental health.

This study affirms the urgent need for government-led initiatives to integrate digital mental health services into Nigeria’s existing maternal healthcare system. Strengthening teletherapy services, enhancing data protection laws, and improving digital literacy among postpartum women are critical steps in ensuring the successful implementation of digital health interventions. Furthermore, fostering collaborations between healthcare providers, technology innovators, and community organizations will enhance the sustainability and impact of these solutions.

In conclusion, leveraging digital health technologies can significantly improve maternal mental health outcomes in Nigeria by expanding access to timely and effective psychological support. However, to maximize impact, stakeholders must address the systemic, cultural, and technological barriers that currently limit adoption. Future research should explore longitudinal assessments of digital mental health interventions to further evaluate their effectiveness and sustainability in different socio-economic and cultural contexts. By prioritizing digital mental health integration within maternal care, Nigeria can take significant strides toward reducing the burden of postpartum depression, preventing maternal suicides, and ultimately improving the overall well-being of mothers and their families.

## Data Availability

All data produced in the present work are contained in the manuscript

## Age Distribution

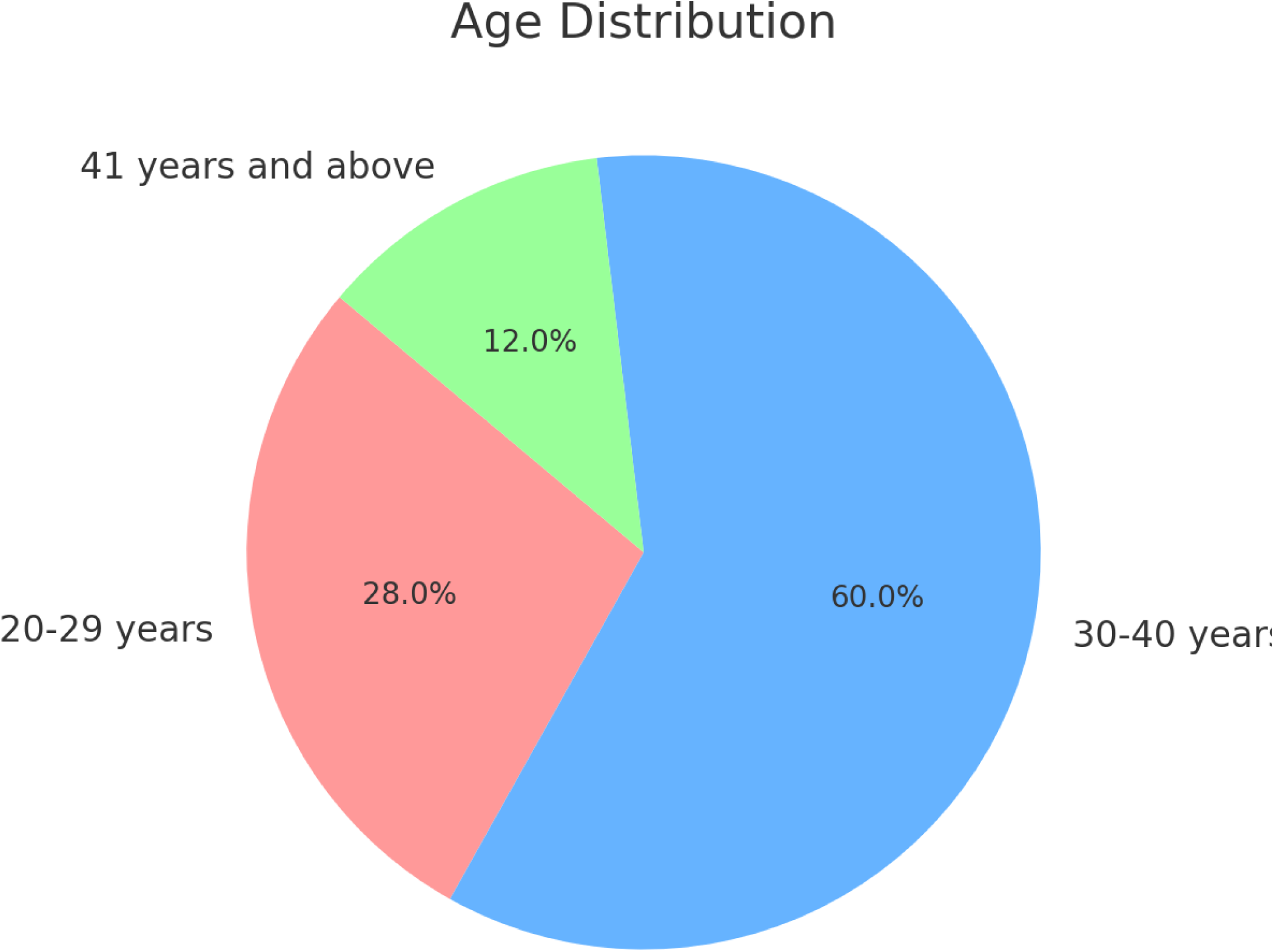

## Educational Level

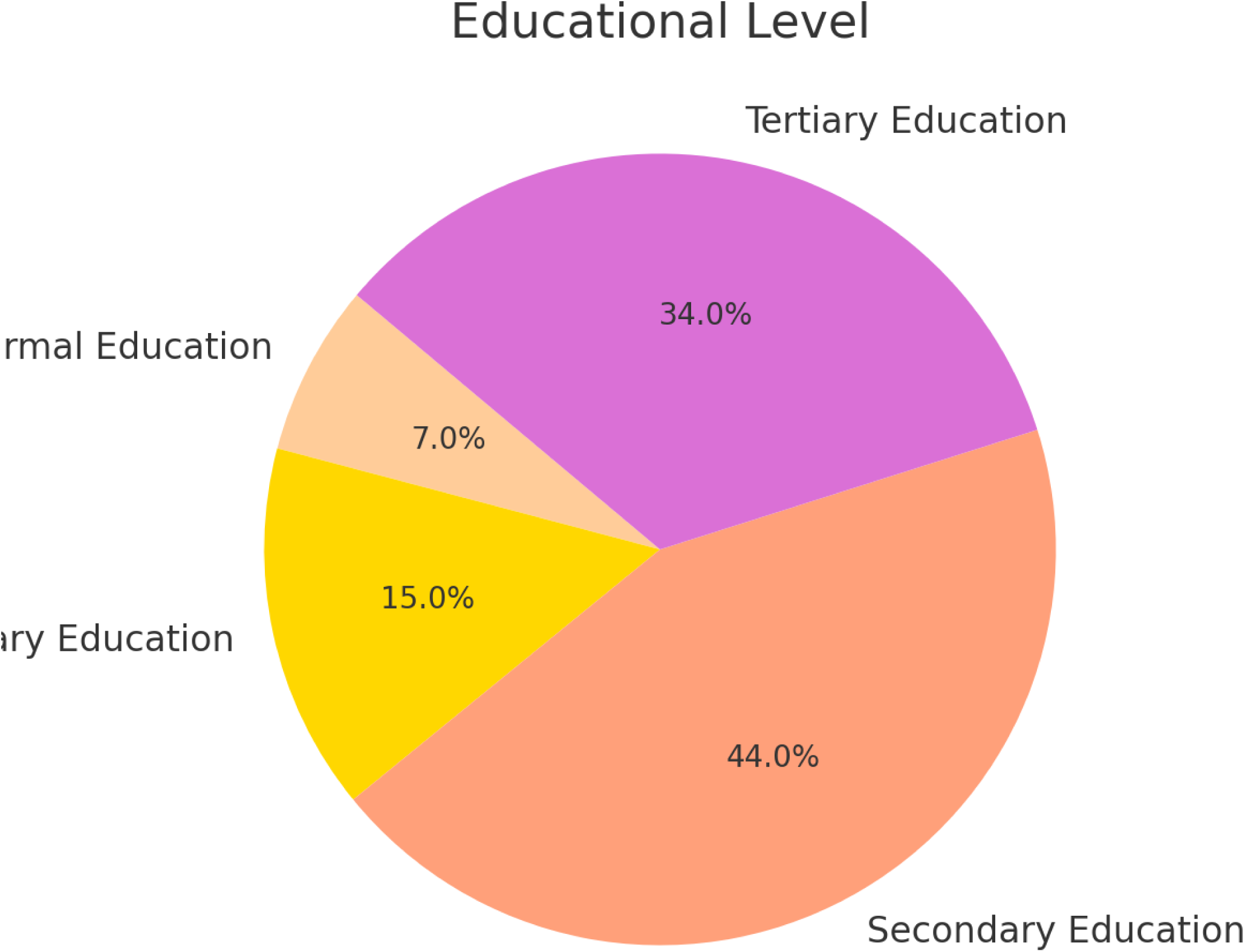

## Employment Status

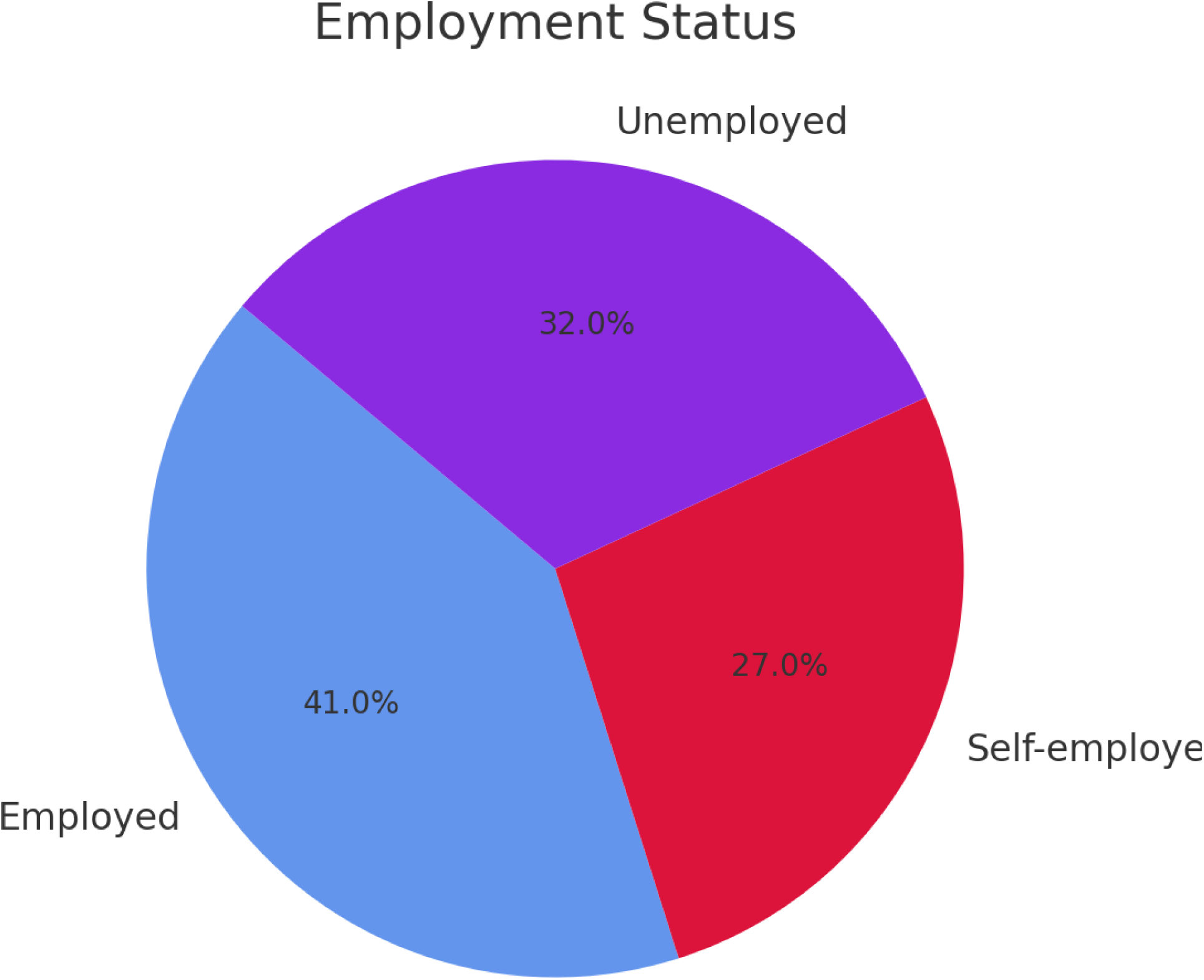

## Household Income Level (Monthly)

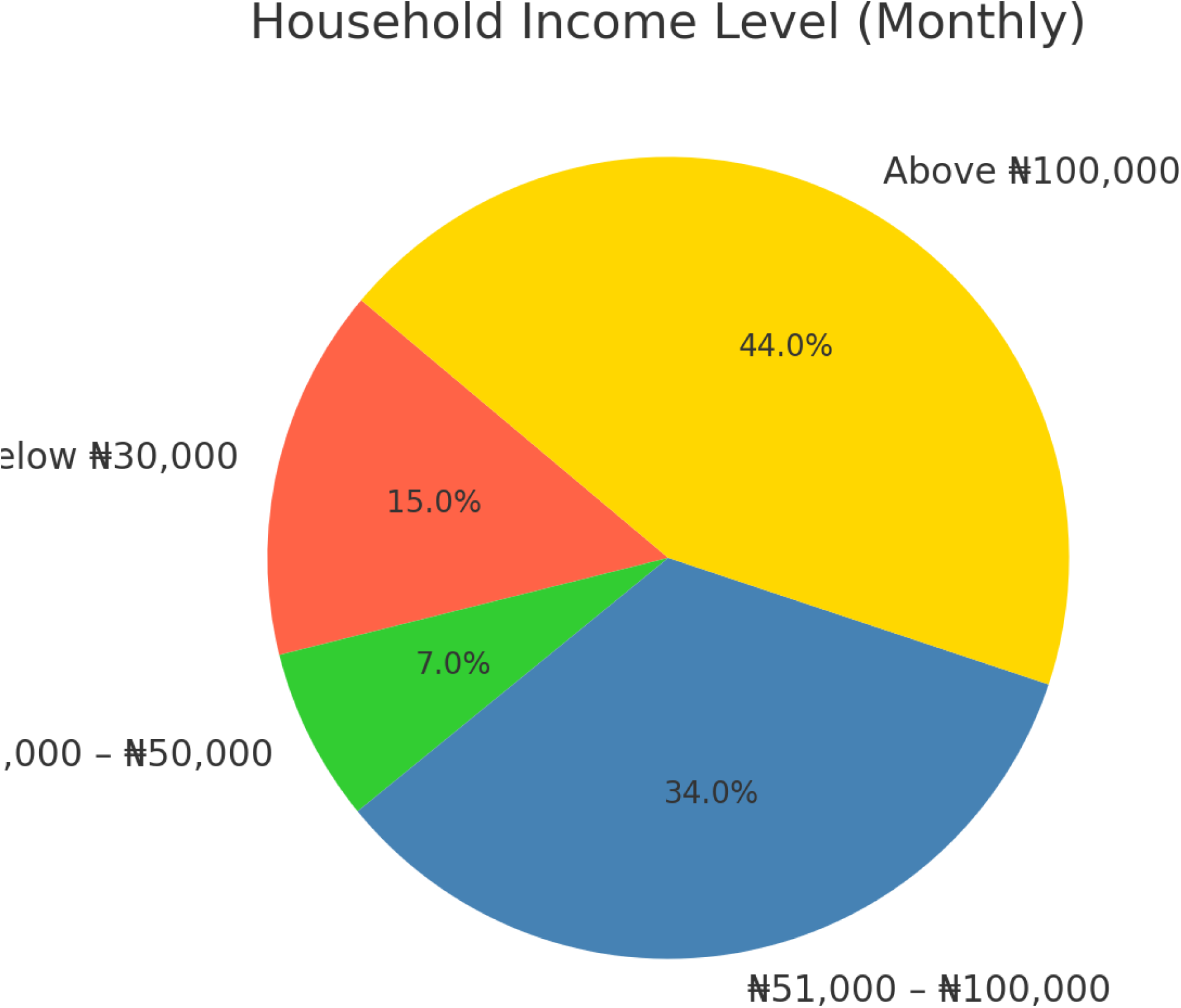

## Marital Status

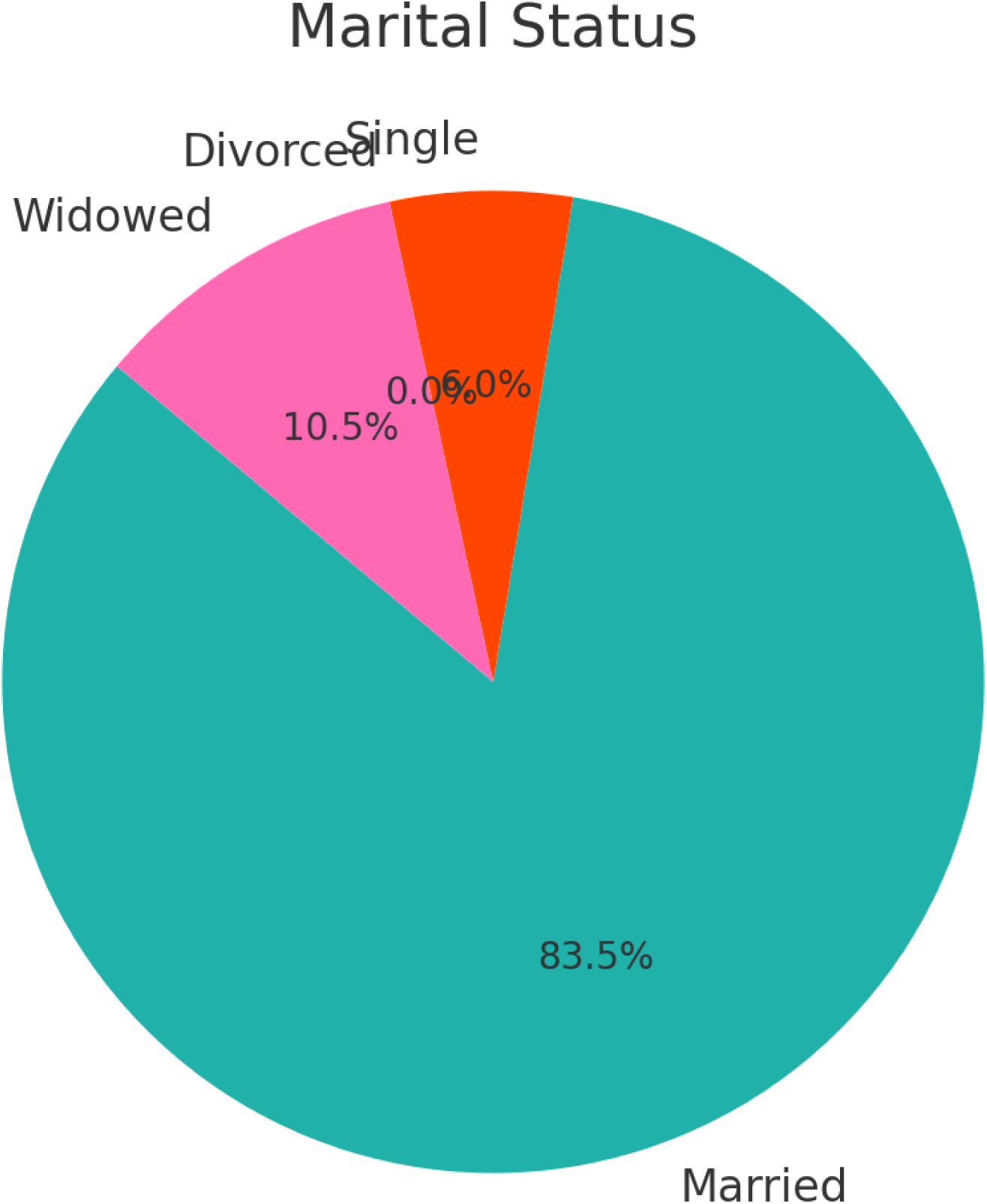

## Feeling Down in the Last Two Weeks

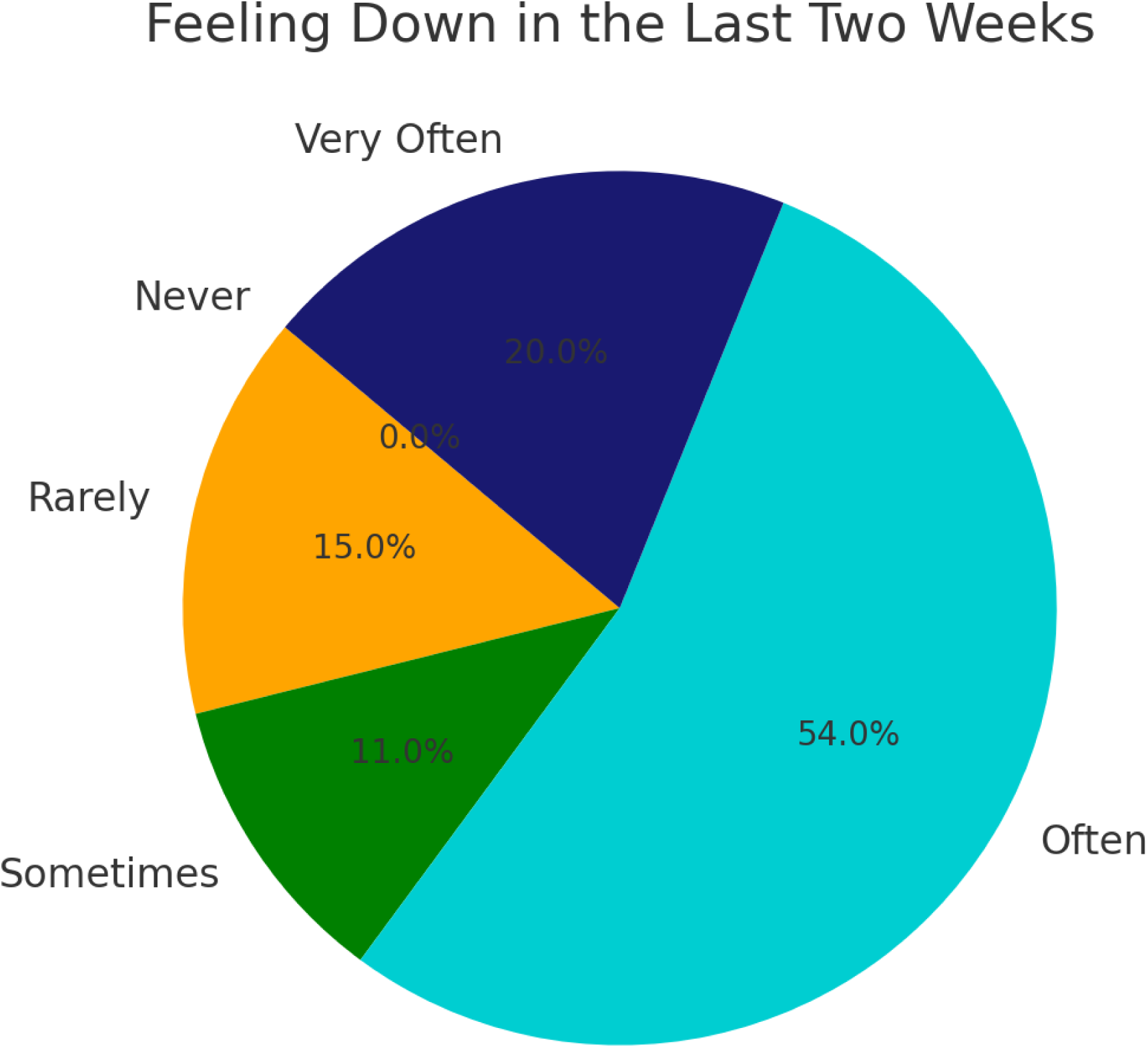

## Received Mental Health Support

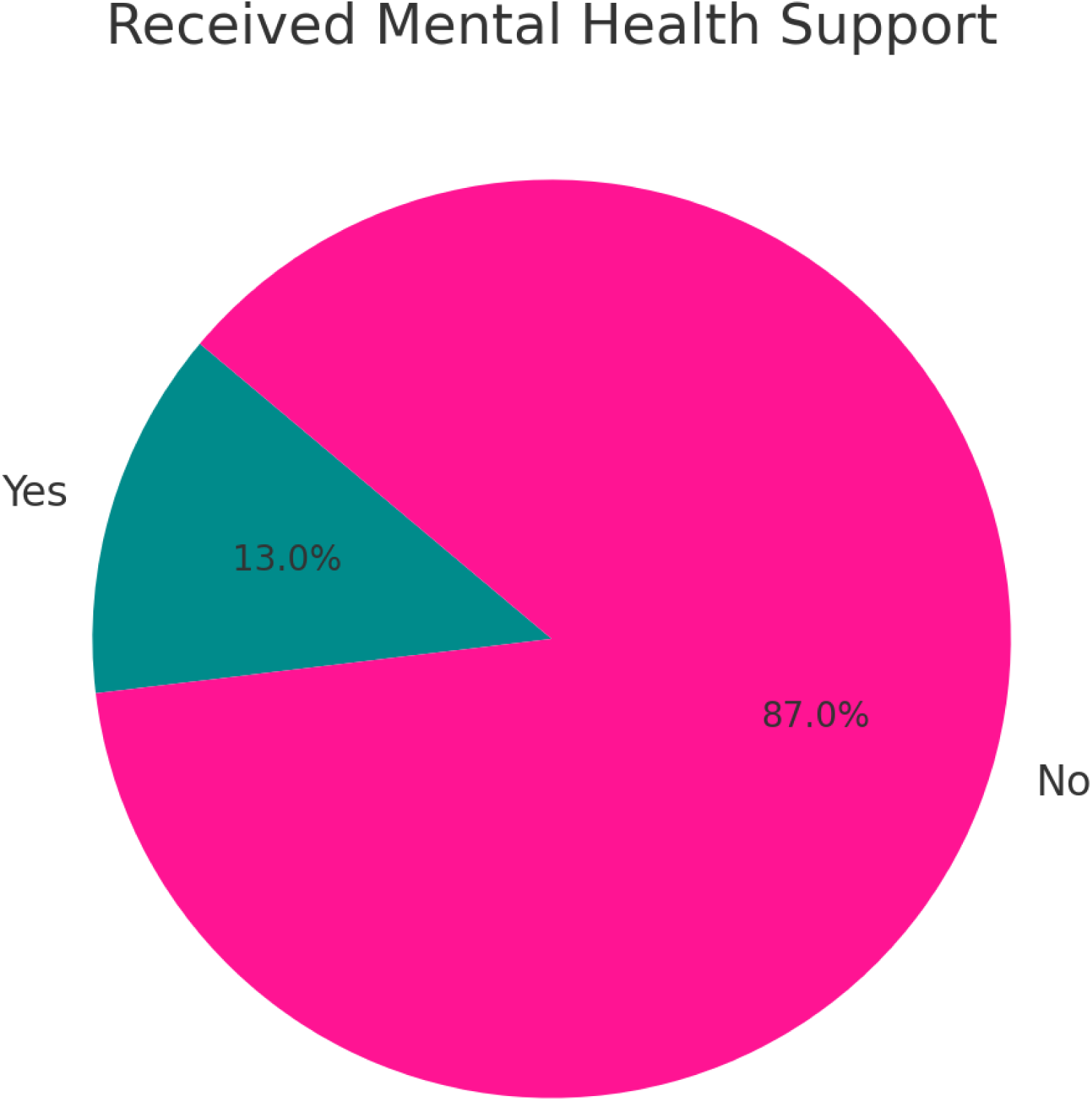

## Moderate to Severe Postpartum Depression

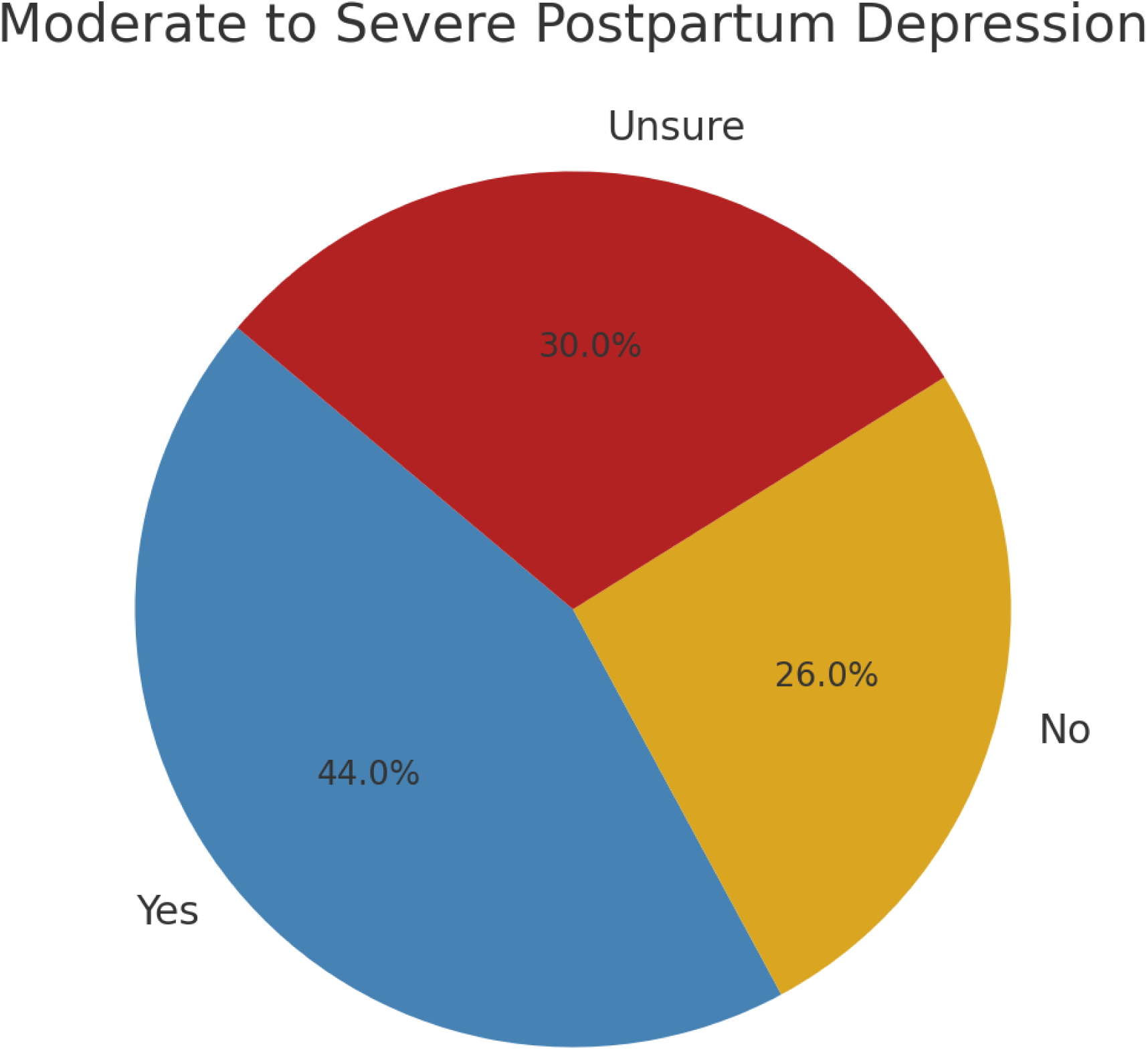

## Spousal Emotional Support

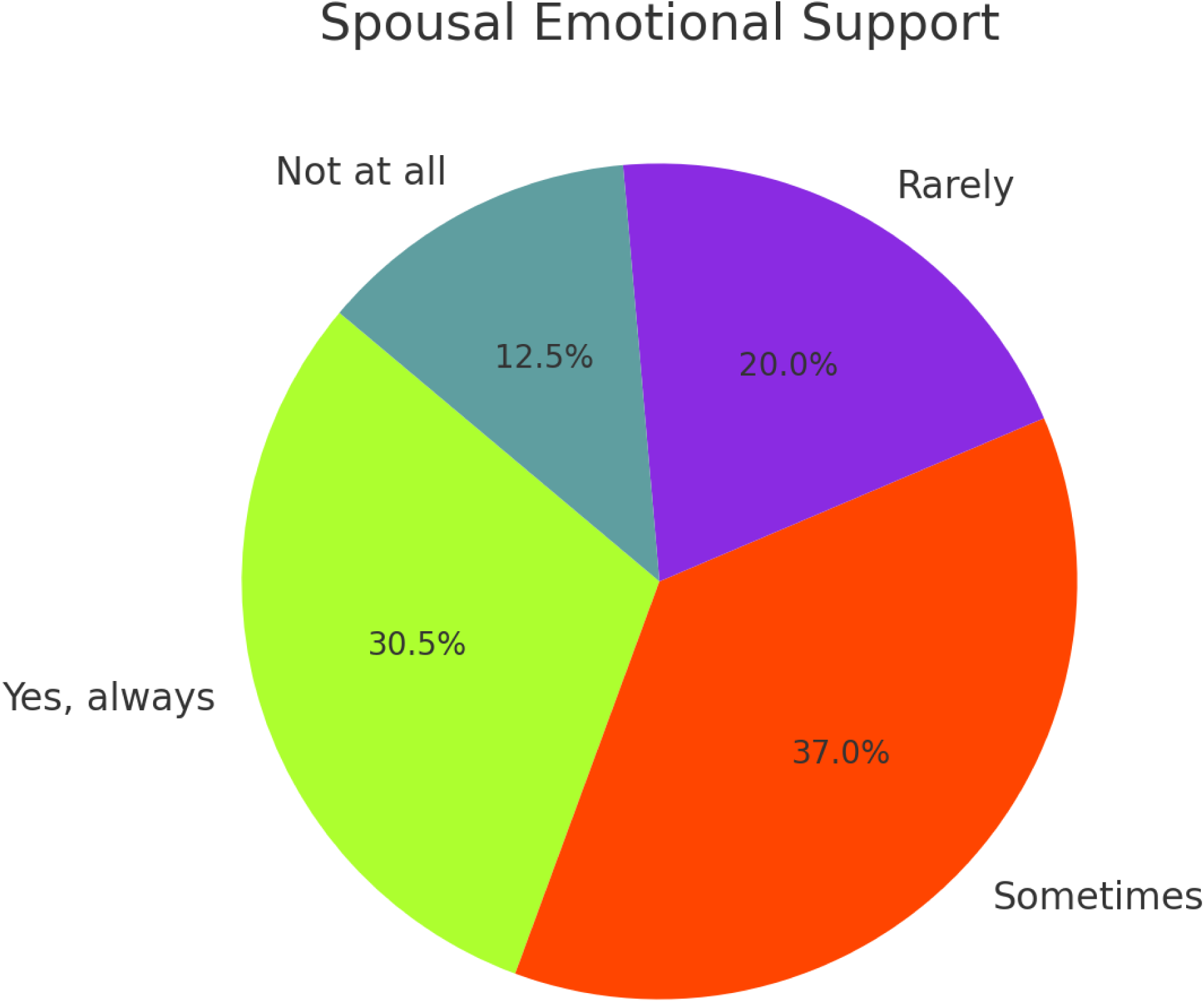

## Comfort in Discussing Emotional Struggles

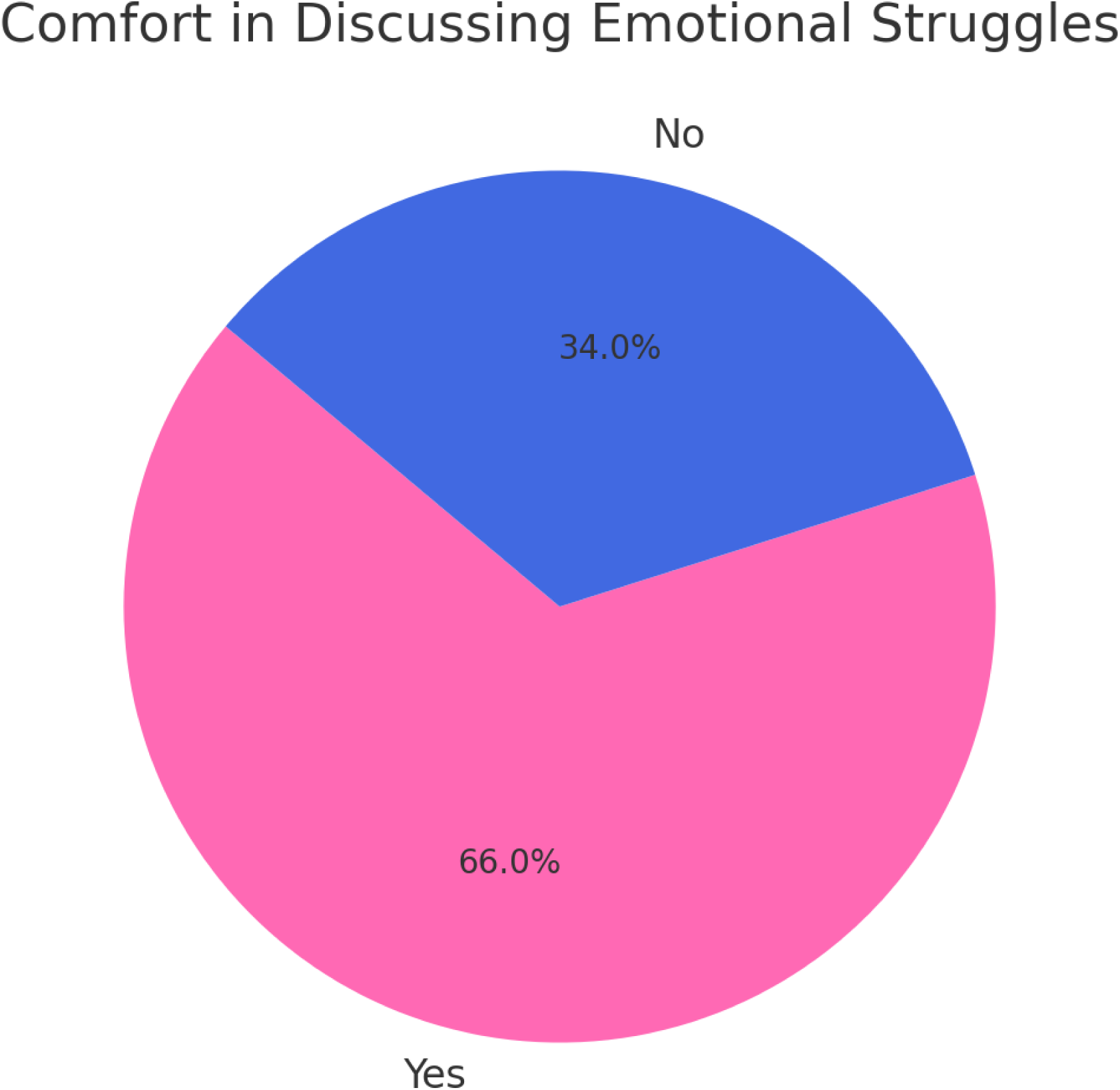

## Used Digital Mental Health Support

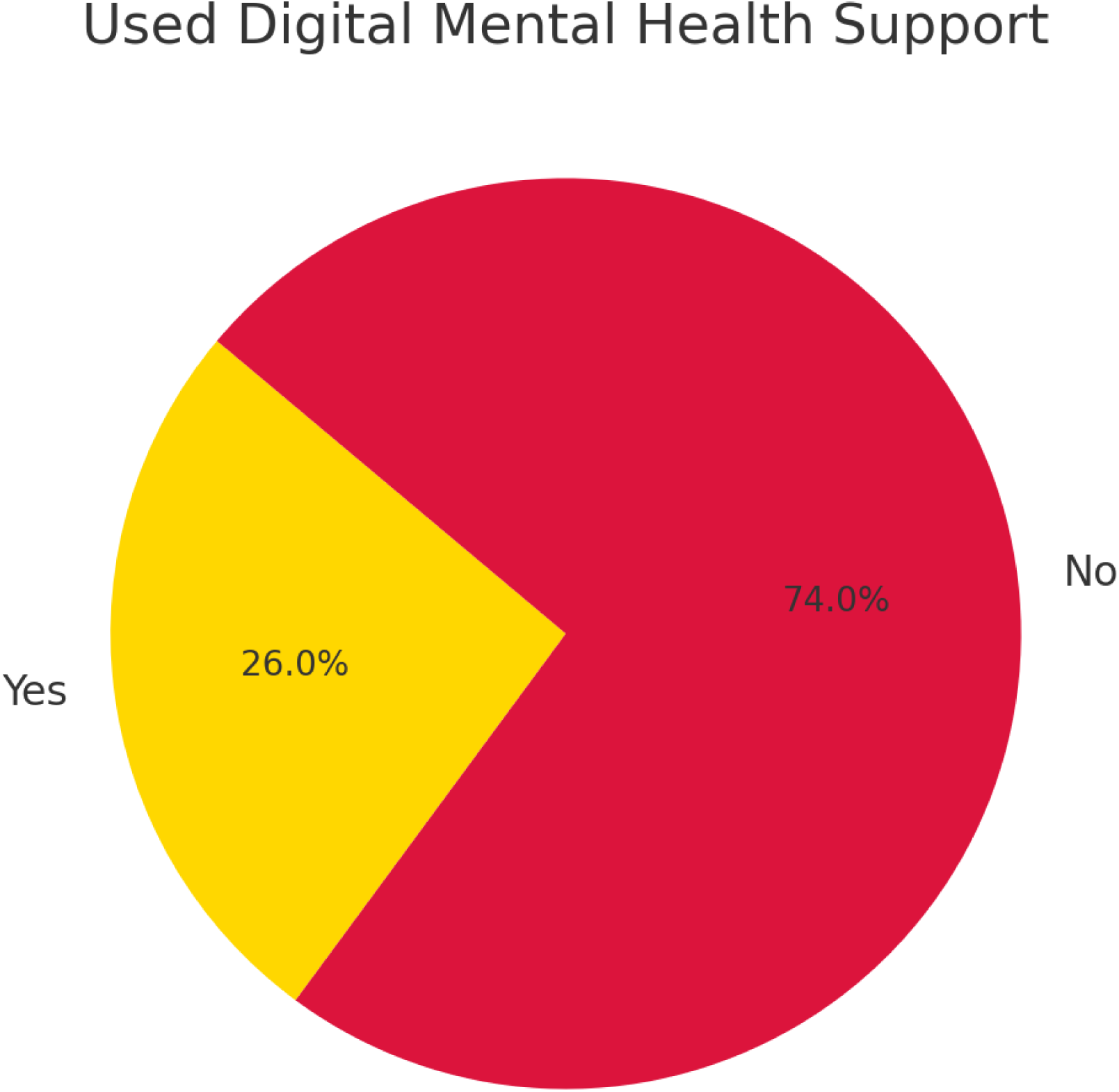

## Willing to Use Digital Mental Health Support

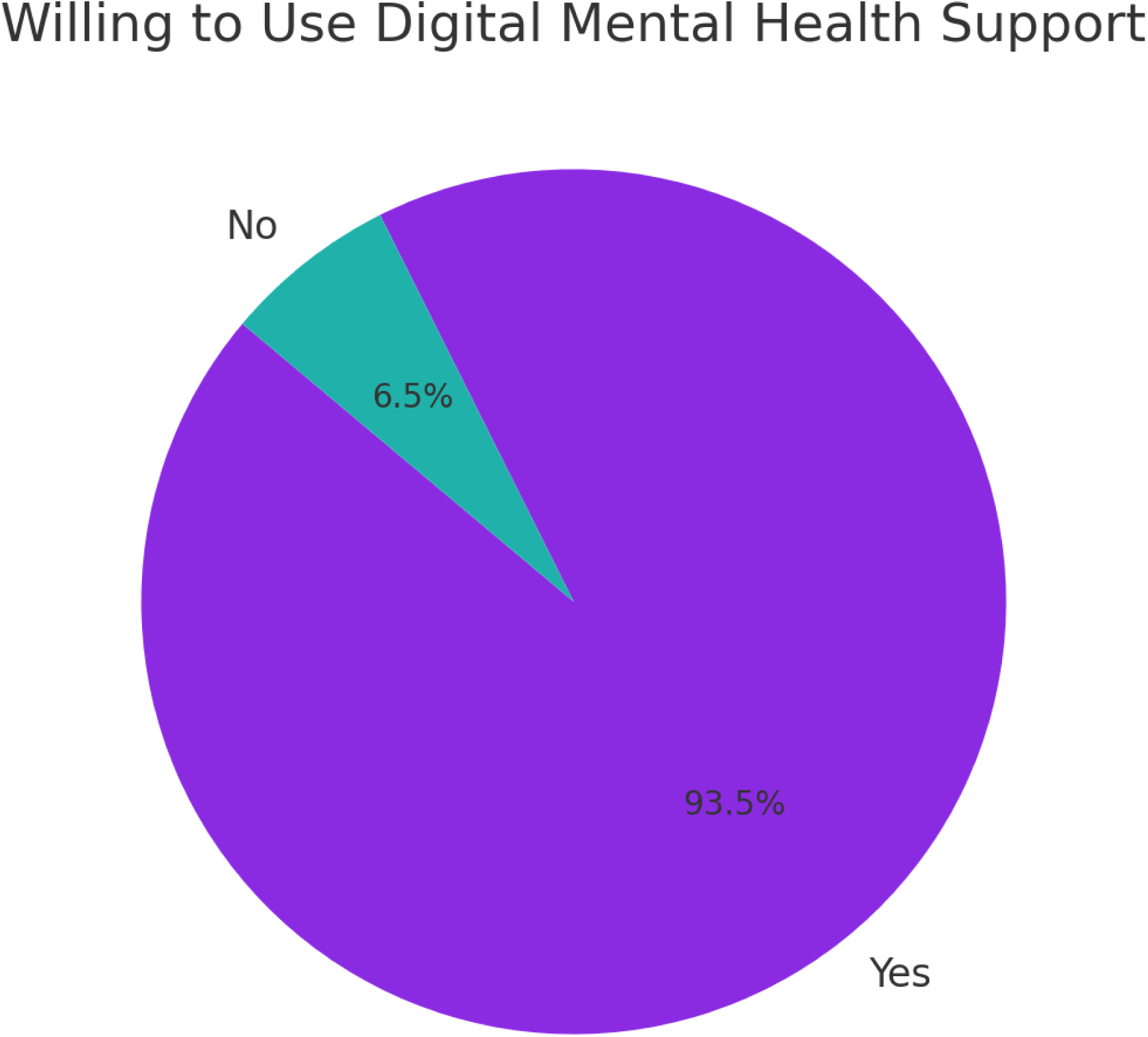

## Survey on Digital Health and Maternal Mental Health in Nigeria

This survey is for postpartum women in Oyo and Ogun State to assess the impact of digital health solutions on postpartum depression and maternal mental health. Please tick (✓) the appropriate answers.

### SECTION A: Demographic Information

1. Age Group

□ 18 – 24 years
□ 25 – 30 years
□ 31 – 35 years
□ 36 – 40 years
□ Above 40 years
2. Education Level:

□ No formal education
□ Primary education
□ Secondary education
□ Higher education
3. Employment Status:

□ Employed (Full-time)
□ Employed (Part-time)
□ Self-employed
□ Unemployed
□ Student
4. Household Income Level (Monthly):

□ Below ₦30,000
□ ₦30,000 – ₦50,000
□ ₦51,000 – ₦100,000
□ Above ₦100,000
5. Marital Status:

□ Married
□ Single
□ Divorced
□ Widowed

### SECTION B: Maternal Mental Health and Postpartum Depression

6. In the past two weeks, how often have you felt down, depressed, or hopeless?

□ Never □ Rarely □ Sometimes □ Often □ Very Often

7. Have you experienced any of the following symptoms since childbirth? (Tick all that apply)

□ Persistent sadness
□ Loss of interest in activities
□ Fatigue or low energy
□ Changes in sleeping patterns
□ Thoughts of self-harm or suicide
8. Have you received any mental health support since childbirth?

□ Yes
□ No
9. If yes, where did you receive support? (Tick all that apply)

□ Hospital/Clinic
□ Religious leader
□ Family/Friends
□ Online counseling
□ Others (Specify)
10. Would you say you have moderate to severe symptoms of postpartum depression?

□ Yes
□ No
□ Unsure

### SECTION C: Risk Factors for Postpartum Depression

11. What challenges have contributed to your emotional struggles after childbirth?** *(Tick all that apply)*

□ Financial stress
□ Lack of spousal support
□ Family pressure
□ Cultural stigma around mental health
□ Physical health complications after childbirth
12. Does your spouse/partner support you emotionally?

□ Yes, always
□ Sometimes
□ Rarely
□ Not at all
13. Do you feel comfortable discussing your emotional struggles with family members?

□ Yes
□ No

### SECTION D: Awareness and Use of Digital Health Solutions

14. Have you ever used a mobile app or online platform for mental health support?

□ Yes
□ No
15. If yes, which digital health solutions have you used? (Tick all that apply)

□ Teletherapy (Online counseling)
□ Mental health chatbots
□ Social media peer support groups
□ Meditation or self-help apps
16. If no, why haven’t you used any digital mental health service? (Tick all that apply)

□ Lack of awareness
□ Internet/data costs
□ Stigma around seeking help
□ Not comfortable with digital solutions

### SECTION E: Willingness to Use Digital Health for Mental Health Support

17. Would you be willing to use a mobile app or online platform for mental health support if it were free or affordable?

□ Yes
□ No
18. What features would you want in a digital mental health platform? (Tick all that apply)

□ Free counseling sessions
□ Anonymous chat support
□ Access to mental health professionals
□ Self-help tools and educational resources

Thank you for your participation! Your responses will remain confidential. ’Femi Banjo (B.Pharm)

Digital Health Researcher

## Survey Results: Digital Health and Maternal Mental Health in Nigeria

Findings from 200 Postpartum Women in Nigeria

1. Demographic Information

Total Respondents: 200 postpartum women across Nigeria

Age Distribution:

- 20-29 years: 56 (28%)
- 30-40 years: 120 (60%)
- 41 years and above: 24 (12%)

Educational Level:

- No Formal Education: 14 (7%)
- Primary Education: 30 (15%)
- Secondary Education: 88 (44%)
- Tertiary Education: 68 (34%)

Employment Status:

- Employed: 82 (41%)
- Self-employed: 54 (27%)
- Unemployed: 64 (32%)

Household Income Level (Monthly):

- Below ₦30,000: 30 (15%)
- ₦30,000 – ₦50,000: 14 (7%)
- ₦51,000 – ₦100,000: 68 (34%)
- Above ₦100,000: 88 (45%)

Marital Status:

- Married 167 (83.5%)
- Single 12 (6%)
- Divorced 0 (0%)
- Widowed 21 (10.5%)

2. Maternal Mental Health and Postpartum Depression

In the past two weeks, how often have you felt down, depressed, or hopeless?

- Never 0 (0%)
- Rarely 30 (15%)
- Sometimes 22 (11%)
- Often 108 (54%)
- Very Often 40 (20%)

Have you received any mental health support since childbirth?

- Yes 26 (13%)
- No 174 (87%)

Would you say you have moderate to severe symptoms of postpartum depression?

- Yes 88 (44%)
- No 52 (26%)
- Unsure 60 (30%)

Risk Factors for Postpartum Depression

Does your spouse/partner support you emotionally?

- Yes, always 61 (30.5%)
- Sometimes 74 (37%)
- Rarely 40 (20%)
- Not at all 25 (12.5%)

Do you feel comfortable discussing your emotional struggles with family members?

- Yes 132 (66%)
- No 68 (34%)

Awareness and Use of Digital Health Solutions

Have you ever used a mobile app or online platform for mental health support?

- Yes 52 (26%)
- No 148 (74%)

Willingness to Use Digital Health for Mental Health Support

Would you be willing to use a mobile app or online platform for mental health support if it were free or affordable?

- Yes 187 (93.5%)
- No 13 (6.5%)

## Conclusion

The survey shows a significant gap in awareness and adoption of digital mental health solutions among postpartum women in Nigeria. While 44% have experienced postpartum mental health issues, only 29% have used digital health tools for support. Major barriers include financial constraints, digital literacy issues, and privacy concerns.

To improve maternal mental health outcomes, it is essential to increase digital health awareness, make teletherapy services more accessible, and address social stigma through community-driven initiatives and government interventions.

## Notes

### Competing Interest Statement

The authors have declared no competing interest.

### Funding Statement

The study did not receive any funding

### Author Declarations

Olabisi Onabanjo University, Faculty of Pharmacy. Faculty of Pharmacy, Ethics Committee

