## Supplemental table for "The Impact of Digital Health on Maternal Mental Health in Nigeria"

Age Distribution

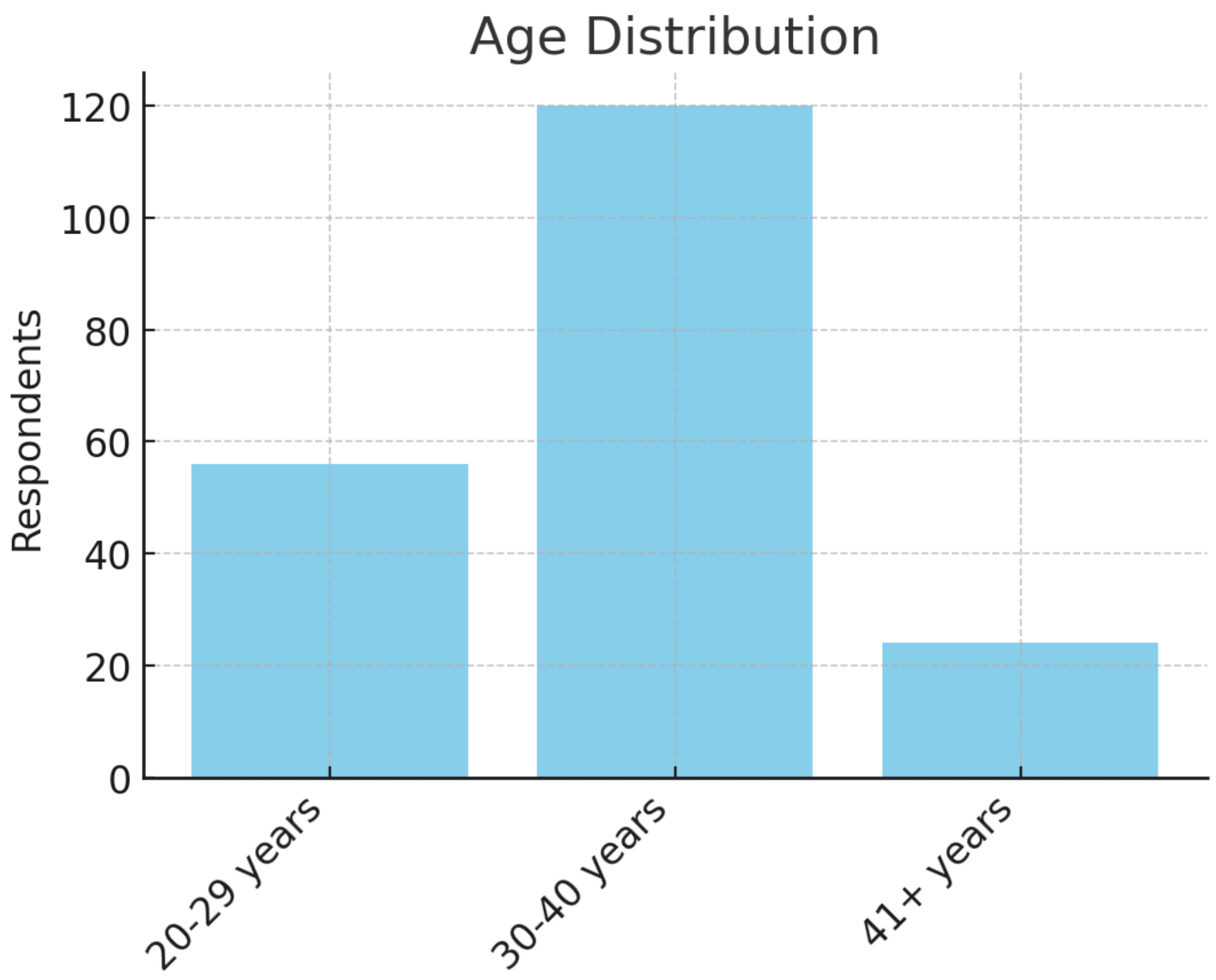

Education Level

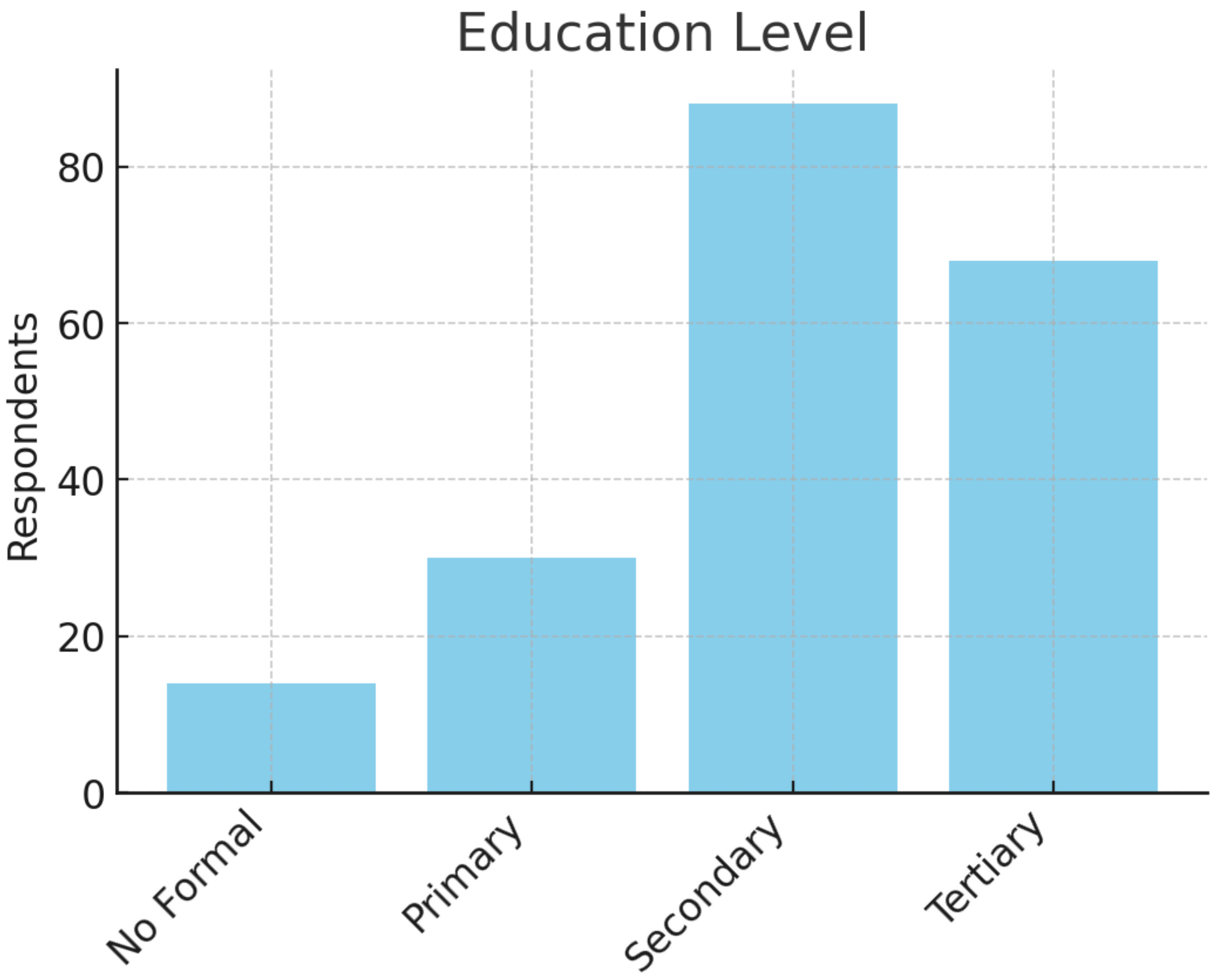

Employment Status

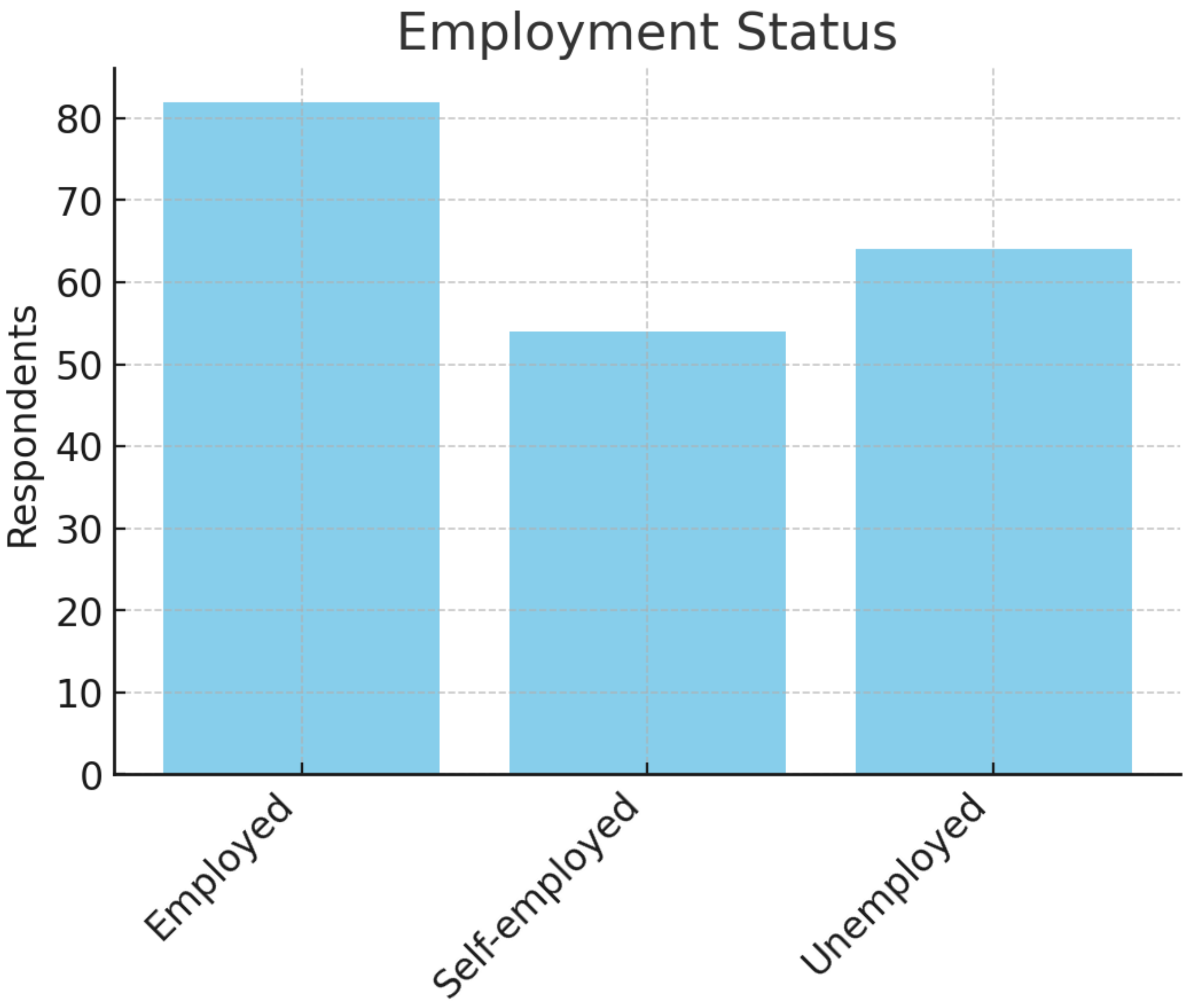

Household Income

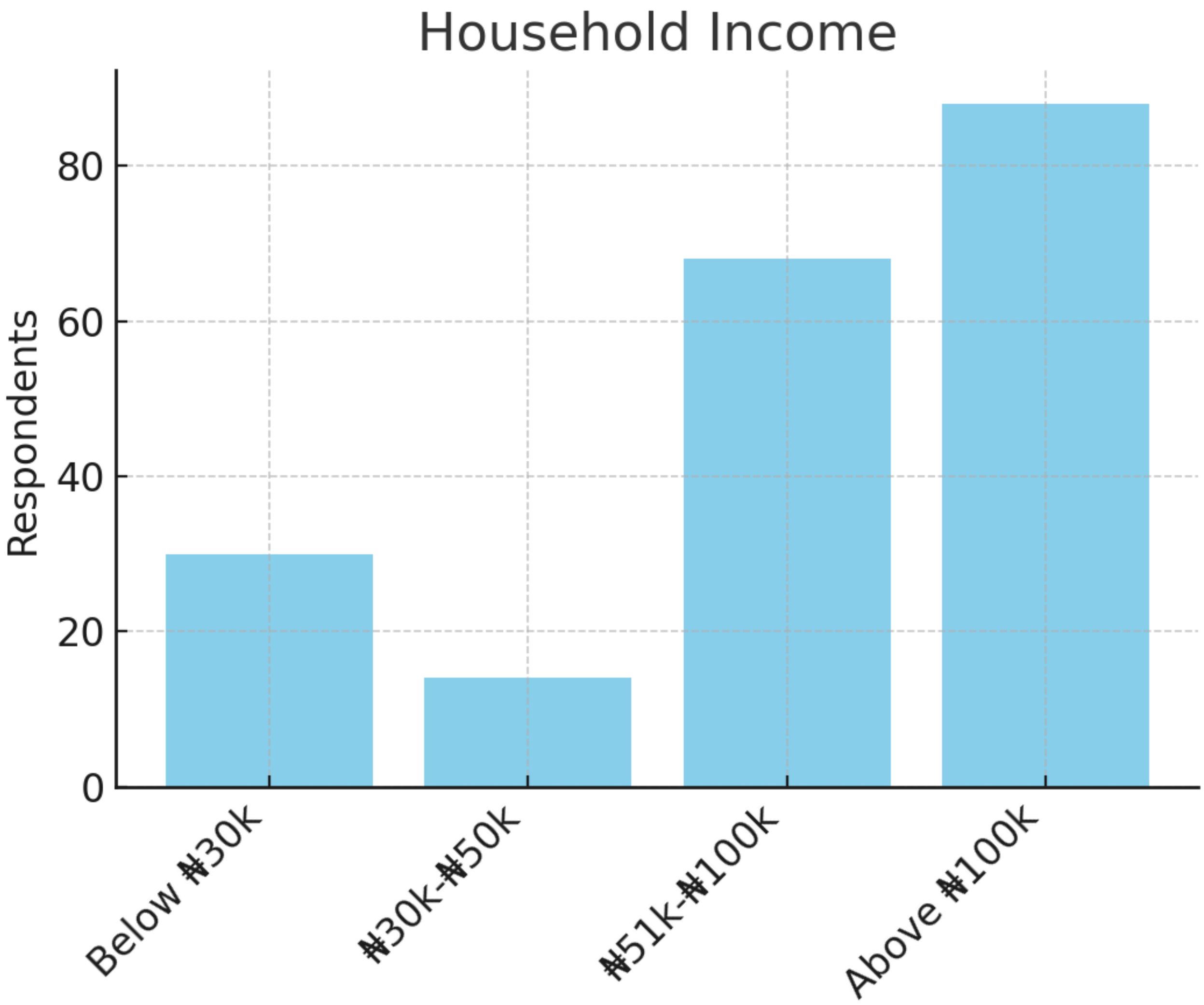

Marital Status

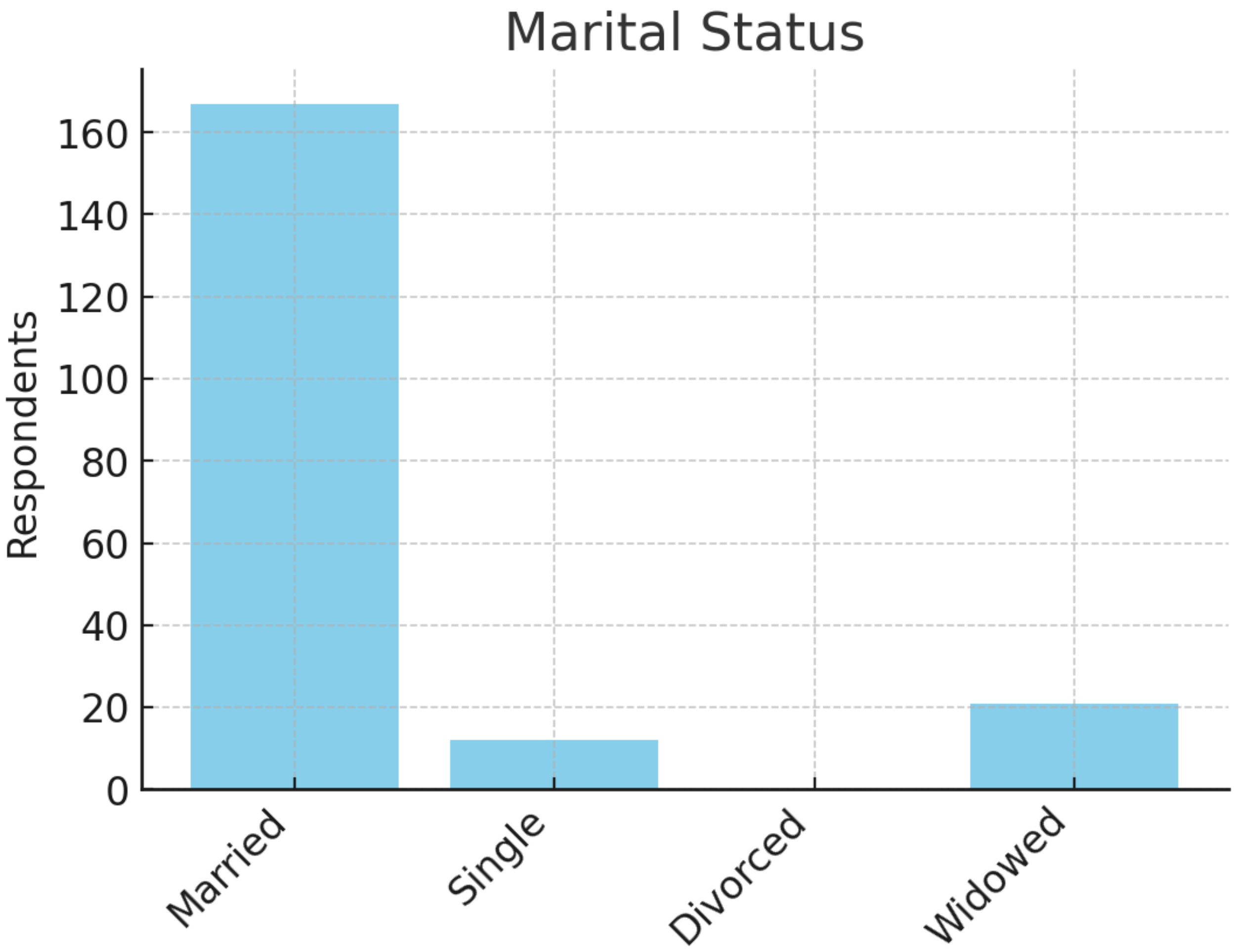

Feeling Depressed

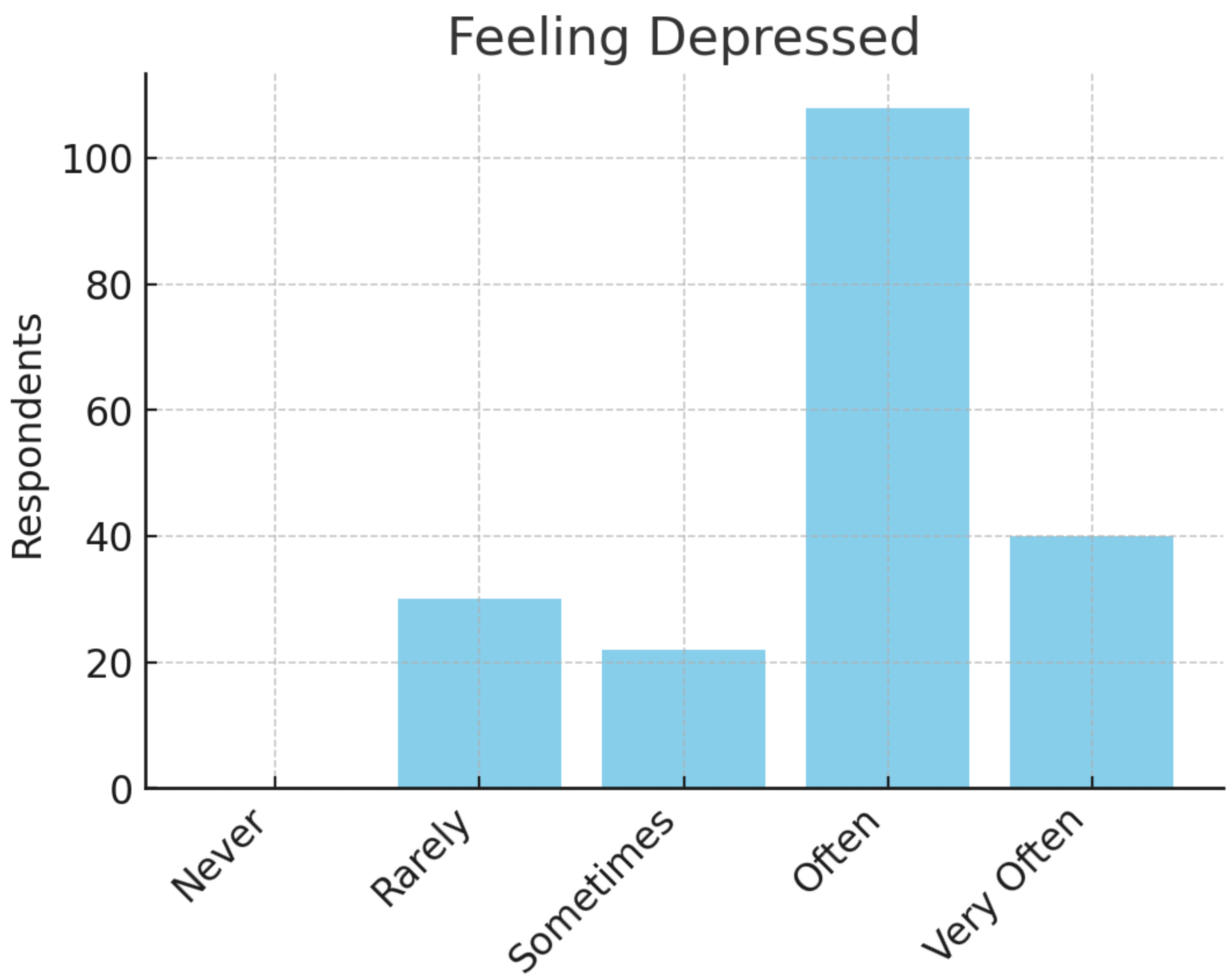

Mental Health Support

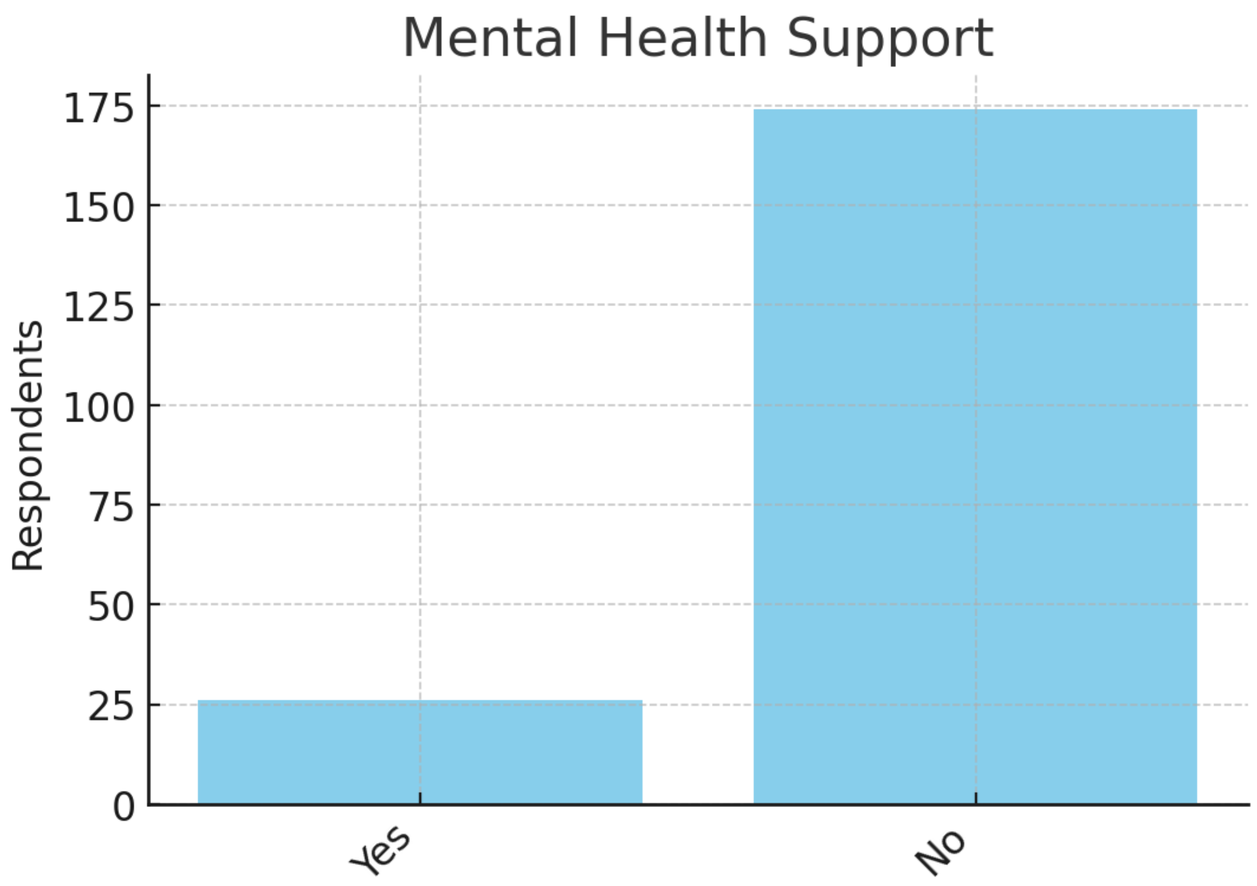

Postpartum Depression

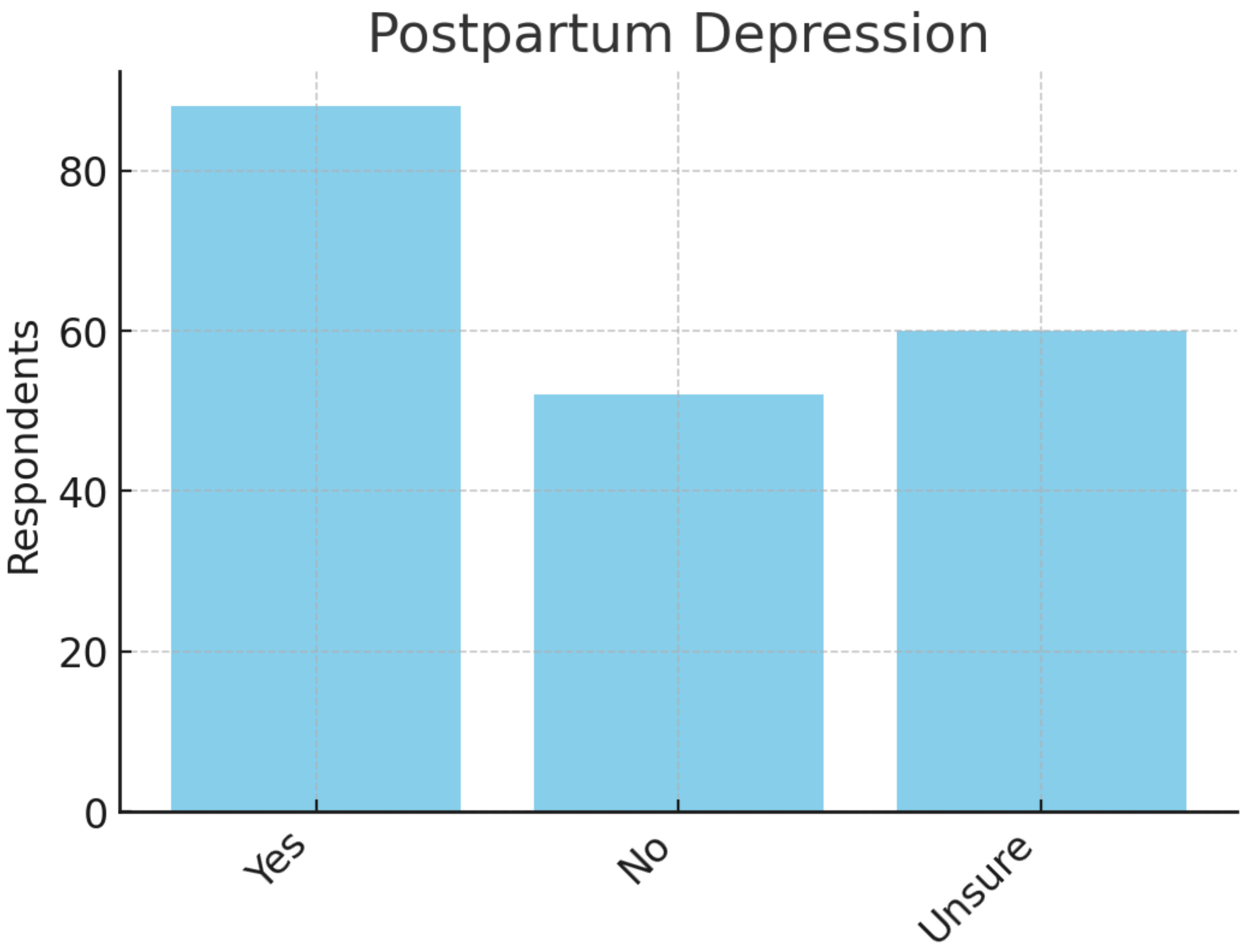

Spousal Support

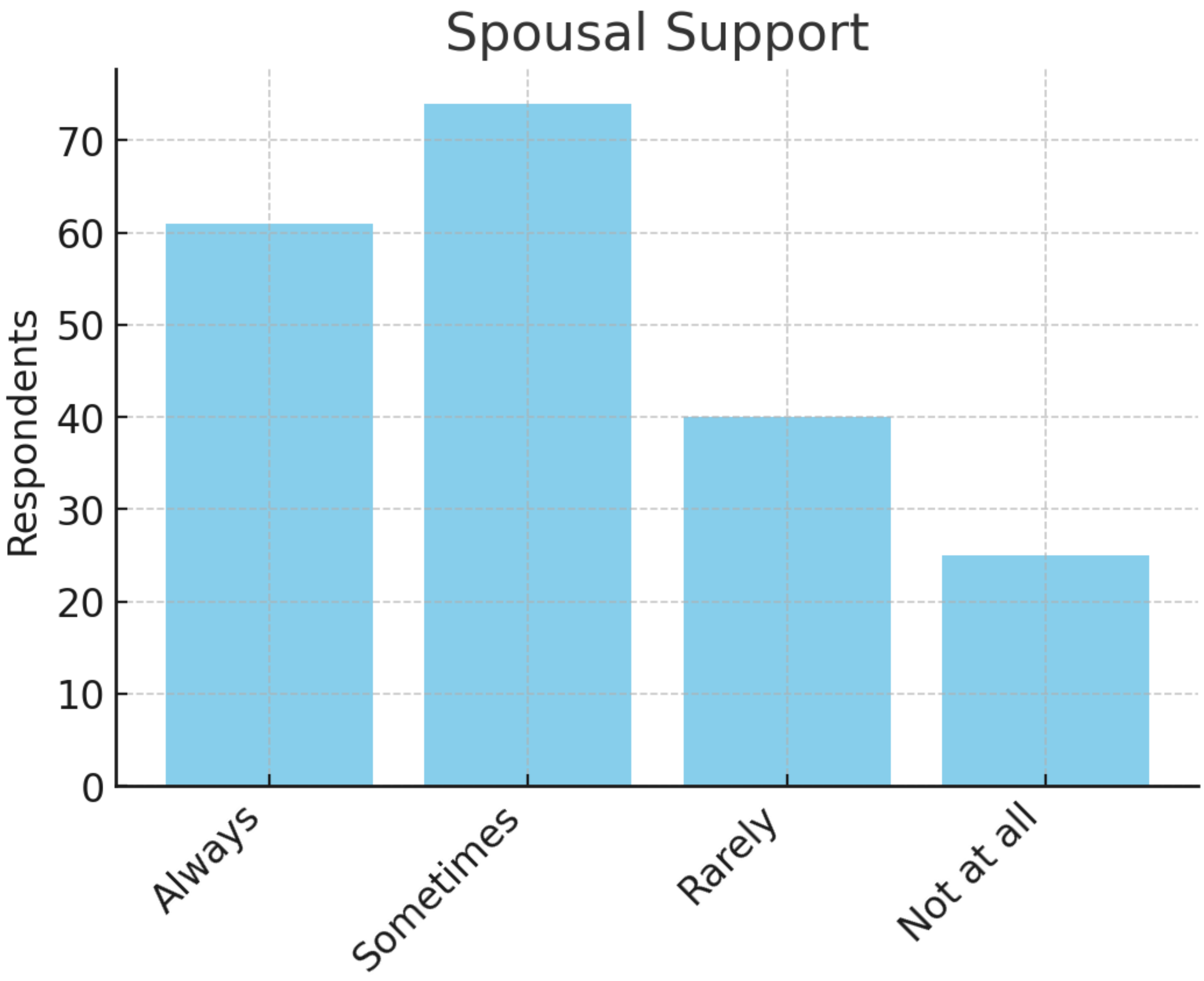

Family Discussion

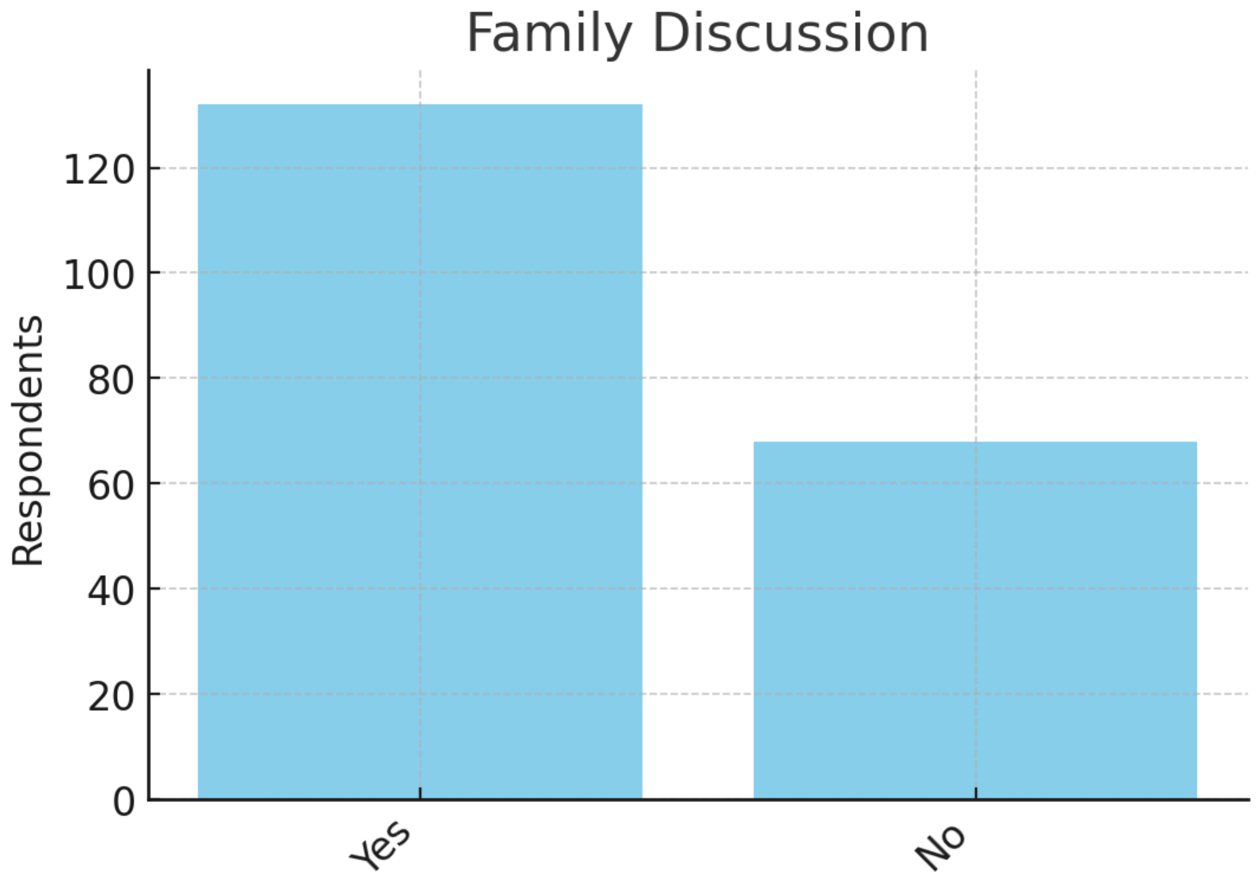

Used Digital Health

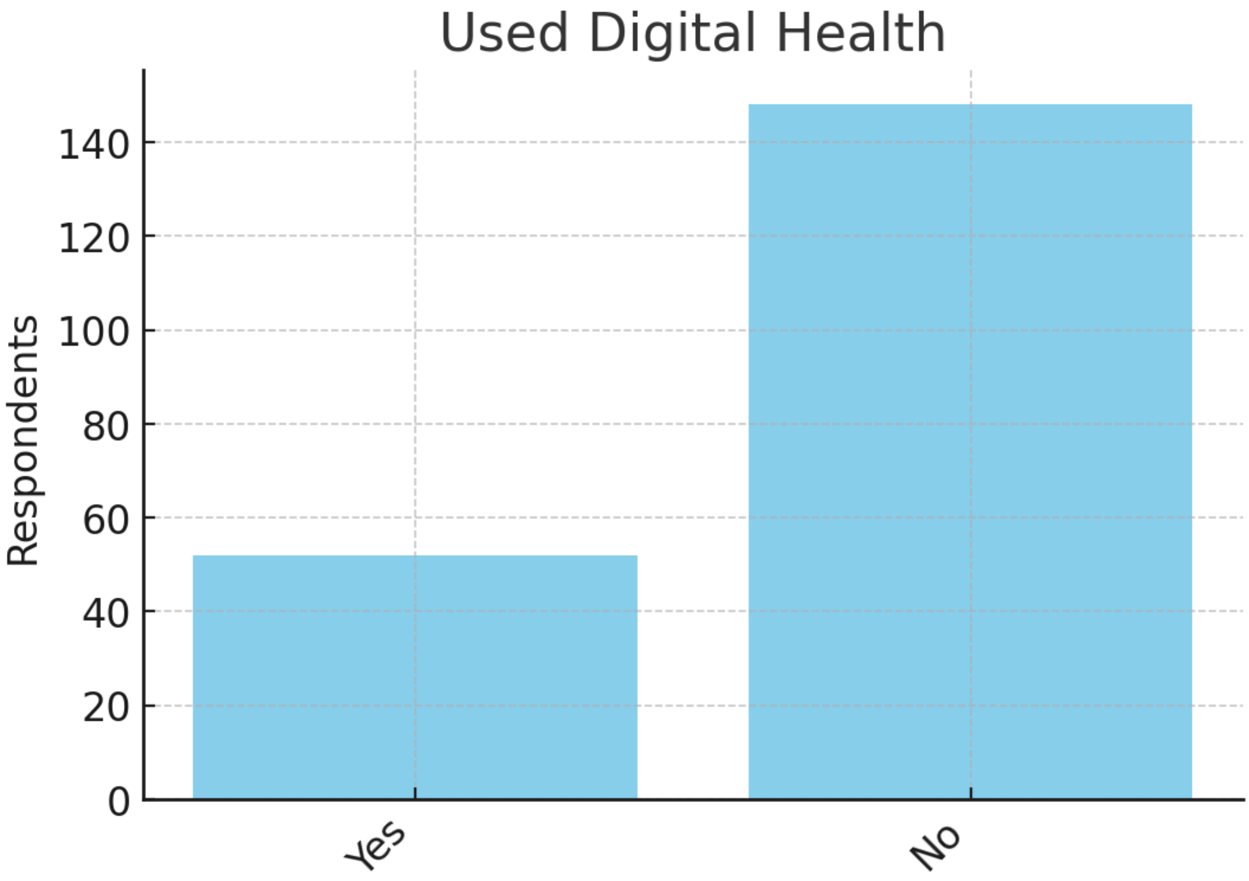

Willing to Use Digital Health

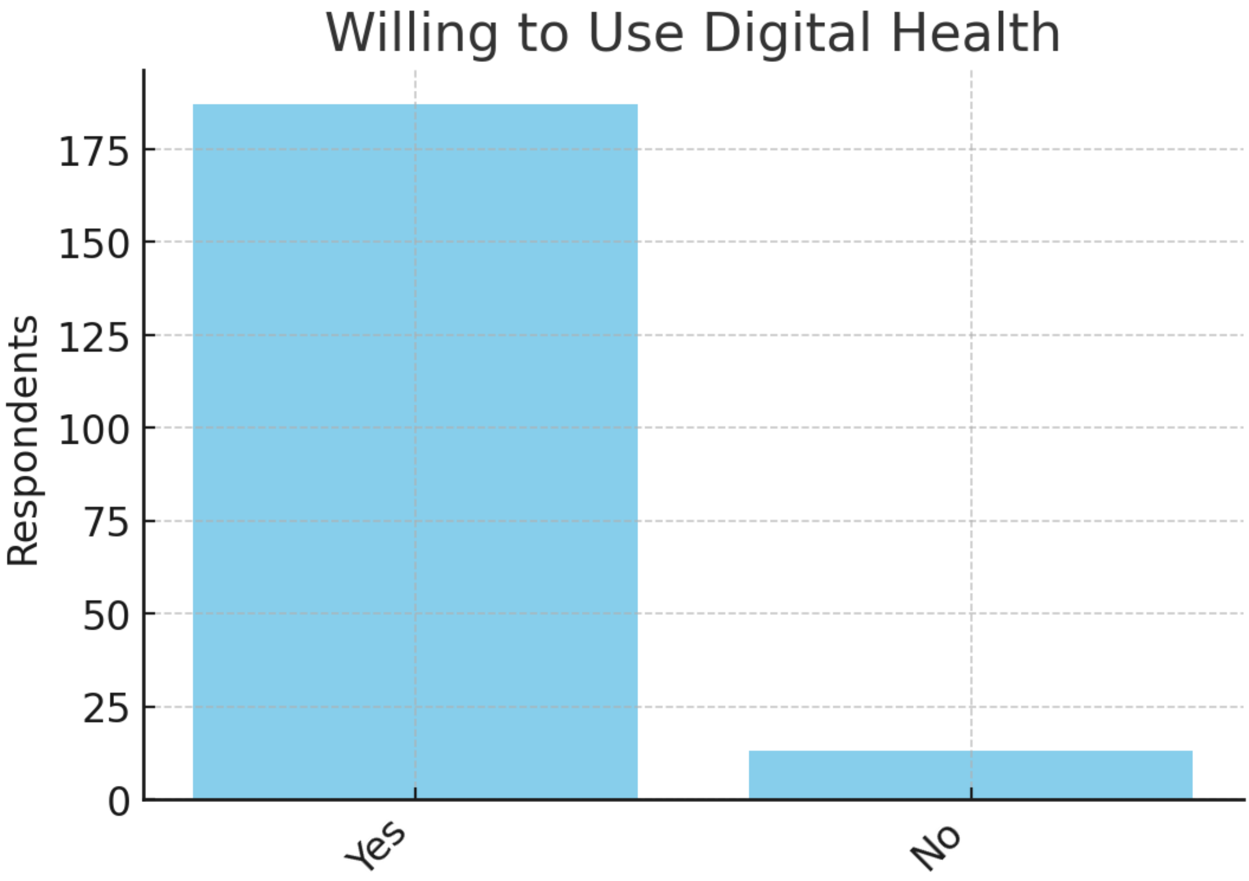
